# Body mass index changes and their association with SARS-CoV-2 infection: a real-world analysis

**DOI:** 10.1101/2024.02.12.24302697

**Authors:** Jithin Sam Varghese, Yi Guo, Mohammed K. Ali, W. Troy Donahoo, Rosette J. Chakkalakal

## Abstract

**Objective:** To study body mass index (BMI) changes among individuals aged 18-99 years with and without SARS-CoV-2 infection.

**Subjects/Methods:** Using real-world data from the One Florida+ Clinical Research Network of the National Patient-Centered Clinical Research Network, we compared changes over time in BMI in an Exposed cohort (positive SARS-CoV-2 test between March 2020 – January 2022), to a contemporary Unexposed cohort (negative SARS-CoV-2 tests), and an age/sex-matched Historical control cohort (March 2018 – January 2020). Body mass index (kg/m^2^) was retrieved from objective measures of height and weight in electronic health records. We used target trial approaches to estimate BMI at baseline and change per 100 days of follow-up for Unexposed and Historical cohorts relative to the Exposed cohort by categories of sex, race-ethnicity, age, and hospitalization status.

**Results:** The study sample consisted of 44,436 (Exposed cohort), 164,118 (Unexposed cohort), and 41,189 (Historical cohort). Cumulatively, 62% were women, 21.5% Non-Hispanic Black, 21.4% Hispanic and 5.6% Non-Hispanic Other. Patients had an average age of 51.9 years (SD: 18.9). At baseline, relative to the Exposed cohort (mean BMI: 29.3 kg/m^2^ [95%CI: 29.0, 29.7]), the Unexposed (–0.07 kg/m^2^ [95%CI; –0.12, –0.01]) and Historical controls (–0.27 kg/m^2^ [95%CI; – 0.34, –0.20]) had lower BMI. Relative to no change in the Exposed over 100 days (0.00 kg/m^2^ [95%CI; –0.03,0.03]), the BMI of those Unexposed decreased (–0.04 kg/m^2^ [95%CI; –0.06, – 0.01]) while the Historical cohort’s BMI increased (+0.03 kg/m^2^ [95%CI;0.00,0.06]). BMI changes were consistent between Exposed and Unexposed cohorts for most population groups, except at start of follow-up period among Males and those 65 years or older, and in changes over 100 days among Males and Hispanics.

**Conclusions:** In a diverse real-world cohort of adults, mean BMI of those with and without SARS-CoV2 infection varied in their trajectories. The mechanisms and implications of weight retention following SARS-CoV-2 infection remain unclear.

## Introduction

Poor cardiometabolic health, including obesity and diabetes, are known risk factors for COVID-19 severity and mortality.^1^ Recent studies of post-acute sequelae of COVID-19 (PASC) suggest COVID-19 may, in turn, worsen cardiometabolic health as increased rates of type 2 diabetes, high blood pressure, and dyslipidemia have been reported for individuals infected with SARS-CoV-2.^2–8^ Few studies have examined changes in body mass index (BMI) which may contribute to observed associations of COVID-19 with incident cardiometabolic diseases, particularly for younger, healthier adults. Racial and ethnic minorities in the US have also been underrepresented in prior studies of PASC, making the findings of this research difficult to generalize to communities that experience disproportionately higher rates of SARS-CoV-2 infection, COVID-19-related morbidity and mortality, and cardiometabolic disease in the US.^9–11^ Understanding the magnitude and direction of changes in cardiometabolic risk factors during the post-acute period of SARS-CoV-2 infection status in diverse, real-world populations can provide insights into pathways through which COVID-19 may increase rates of type 2 diabetes and other cardiometabolic conditions.

The purpose of this real-world evidence study was to characterize key changes in BMI among individuals who survived the acute phase (defined as the first 30 days) of SARS-CoV-2 infection in a socio-demographically diverse cohort and compare them with a contemporary cohort who always tested negative for SARS-CoV-2 as well as a historical cohort (prior to the pandemic) matched on age and sex to those who tested positive. Using analytical approaches for target trial emulation, we examined if BMI changes differed by socio-demographic (sex, race-ethnicity, age) category and hospitalization status.

## Methods

### Study Design

We conducted a secondary analysis of patient-level electronic health record (EHR) data from the OneFlorida+ clinical research network (CRN) in the National Patient-Centered Clinical Research Network (PCORnet).^12^ PCORnet CRNs have become a valuable resource to study symptom profiles of PASC.^13,14^ For the current study, the OneFlorida+ Data Analytics team independently queried the clinical data warehouse, built on the PCORnet Common Data Model, to extract data on socio-demographic and clinical characteristics of eligible patients.^15^

We built three retrospective cohorts of adult individuals who sought care at least three times in the OneFlorida+ clinical sites over the period 2016-22.^12^ Individuals were included if they were 18 years or older with at least two inpatient or outpatient encounters recorded in OneFlorida+ in the two years before inclusion, and at least one inpatient or outpatient encounter in the period after inclusion (30 days from inclusion until last visit or 28 February 2022). Individuals were excluded if they had diabetes mellitus, as defined by previously validated computable phenotypes for type 1 diabetes (T1DM) and type 2 diabetes (T2DM; at least two of three criteria: ICD-9/10 diagnosis code for diabetes, antidiabetic medication, and/or HbA1c ≥6.5%), in the 24 months preceding the date of first test or COVID-19 diagnosis code (**Supplementary Table 1)**.^16^

The Institutional Review Board of Emory University determined the study met criteria for exemption under 45 CFR 46.104(d). This study followed the REporting of studies Conducted using Observational Routinely-collected health Data (RECORD) Statement.^17^

### Exposure Cohorts

The Exposed cohort were individuals exposed to SARS-CoV-2, defined by meeting at least one of two criteria between 1 March 2020 and 29 January 2022: any positive test (Nucleic Acid Amplification Tests [NAAT] or antigen) or COVID-19 related ICD-10-CM codes (E08 to E13).^13^ The index date of infection exposure (T_0_) was defined as the date of the first positive test or date of record for the ICD-10-CM code. The timeline for cohort selection and follow-up is presented in **Supplementary** Figure 1.

The Unexposed cohort were individuals not measured to have had exposure to SARS-CoV-2 and consisted of individuals who met all of three criteria between 1 March 2020 and 29 January 2022: at least one negative test (NAAT or antigen), no COVID-19 related International Classification of Diseases-10 CM (ICD-10 CM) codes (E08.X to E13.X), and no positive test during follow-up period. Since the Unexposed cohort tested negative throughout the follow-up period, to maximize follow-up duration, the index date (T_0_) was defined as the date of the first negative test.

The Historical cohort was constructed to be representative of typical clinic visits among patients who utilized the healthcare delivery systems in the pre-pandemic period, and ensure differences observed in the Exposed and Unexposed cohorts were not due to the pandemic period. Patients who were not subsequently part of Exposed or Unexposed cohorts were sampled into the Historical cohort after matching on sex and 5-year age intervals. The index dates of Historical cohort ranged from 2 March 2018 and 30 January 2020, and were selected such that the last date of possible follow-up (29 February 2020) did not overlap with the start of the calendar period of identification (1 March 2020) for the Exposed and Unexposed cohorts.

We restricted our study sample to individuals who had at least one measurement of weight in the year preceding their observation period in our study (**Supplementary** Figure 2). The final analytic sample consisted of 44,436 Exposed, 164,118 Unexposed controls and 41,189 Historical controls. All subsequent analysis accounted for the stratified sampling of Historical controls.

### Outcomes – Cardiometabolic Health Indicators

We explored average longitudinal change in body mass index (BMI) based on objective measures recorded in the EHR. Follow-up period (in days) was calculated as number of days from 30 days after index date (i.e. Time _tF_ = t – [T_0_ + 30]), defined as the post-acute phase of the disease. To exclude weight changes associated with severe illness and weight loss prior to mortality, we included only those participants who were alive at least 30 days after SARS-CoV2 infection was detected.^2,13^

### Effect Modifiers – Socio-demographic Characteristics and Hospitalization Status

Socio-demographic characteristics included sex (male, female), race-ethnicity (Non-Hispanic White, Non-Hispanic Black, Hispanic), and age at index date (18-39 years, 40-64 years, 65 years and older). We determined all-cause hospitalization status (not hospitalized, hospitalized) based on the clinical encounter coded on the index date.

### Confounders

We used a combination of biologically plausible covariates and empirically identified covariates to adjust for baseline differences between cohorts using inverse probability weighting (IPW) (see **Supplementary Methods**). Biologically plausible covariates known to be associated with higher cardiometabolic risk were derived from data collected before the index date. We used data collected within 1 year of index date to define smoking status (yes/no), use of medications relevant to cardiometabolic health (yes/no; antihypertensives, statins, antidepressants, antipsychotics, steroids), and laboratory parameters (HbA1c, random glucose, lipid panel). Given the lower frequency of reporting diagnosis codes, we used data collected within 2 years before the index date to identify comorbidities using grouped ICD-10-CM codes (yes/no; obesity, cardiovascular disease, cerebrovascular disease, hyperlipidemia, hypertension, pulmonary disease). Information on medication, laboratory, and diagnosis codes used for categorization are provided in **Supplementary Table 1**. We identified the health system and primary insurance type (Medicare, Medicaid, Other Government, Private, No Insurance, No Information) used on index date.

Empirically identified covariates were computed across six data dimensions available in OneFlorida+: inpatient diagnoses, outpatient diagnoses, inpatient procedures, outpatient procedures, prescriptions, and laboratory tests.^18^ We used participant records from within 1 year before index date to compute the covariates. Diagnostic codes and prescriptions were categorized into 537 Clinical Classifications Software Refined (CCSR) categories and 89 Anatomical Therapeutic Chemical (ATC) Level 3 classes respectively. The 1,951 procedure codes (Current Procedural Terminology, ICD-10-CM) and abnormalities in 876 laboratory tests (Logical Observation Identifiers Names and Codes [LOINC]) were included without additional categorization. We identified the top 200 covariates in each data domain based on their prevalence in the sample. For each covariate, we then computed three intensity dummy variables (at least once, sporadic [more often than median participant], frequent [more often than the 75^th^ percentile]).

### Statistical Analysis

We present an overview of the analytic strategy in **Supplementary** Figure 3.

#### Inverse probability weighting for cohort membership and follow-up

Since this is an observational study, baseline characteristics may differ between exposed, unexposed, and historical control cohorts resulting in non-random cohort membership. To minimize instrumental variable bias when constructing propensity scores, we performed variable selection of empirically identified covariates longitudinally associated with BMI after adjusting for multiple comparison correction using the Benjamini-Hochberg procedure. Next, random forests models for probability of membership in each exposure cohort, relative to other cohorts, were fit separately with 5-fold cross-validation. We tuned the parameters based on the area under receiver operating characteristic curve (AUC) and identified 2000 trees and 10 observations per node as the best combination of hyperparameters. We included both biologically-plausible and algorithmically selected covariates to estimate the high-dimensional propensity score.^19,20^. We assessed covariate balance between the Unexposed and Historical Control cohorts, relative to the Exposed, after IPW using population standardized bias.^21^ Population standardized bias is the maximum difference between IP weighted group mean and unweighted pooled mean.^21^ Bias less than 0.1 after weighting was considered as indicative of covariate balance.

We additionally constructed IPW for availability of follow-up data for BMI to minimize selection bias, since not all participants had data on all cardiometabolic health indicators during the follow-up period.^22^ The final weight for each individual in the analytic sample is a product of treatment weights and selection weights. We provide additional detail of the modeling strategy in an extended methodological note (**Supplementary Methods)**.

IPW were also constructed separately for each socio-demographic, clinical, and community characteristic of interest with numerator reflecting probability of cohort membership under levels of each characteristic (sex, race-ethnicity, age) when assessing differences between levels.^23^

#### Changes in BMI

First, we modeled BMI in the follow-up period for Exposed, Unexposed, and Historical Control cohorts using marginal structural models. We used a difference-in-differences framework to understand if changes in BMI per 100 days of follow-up differed for Unexposed and Historical cohorts relative to the Exposed cohort. We adjusted for all imbalanced characteristics except laboratory parameters in the regression model since the latter had high rates of missingness. Second, we estimated average BMI at origin date and longitudinal change in BMI per 100 days of follow-up across categories of sex (male vs female), race-ethnicity (NH White vs NH Black vs Hispanic), age (18-39 vs 40-64 vs 65 and older) and hospitalization status by exposure group. We did not adjust for multiple comparisons in these prespecified subgroup analysis.

### Sensitivity Analysis

First, to minimize the influence of patients who rarely used the health system, we restricted the analytic sample to those patients with at least 100 days of follow-up and at least two encounters where BMI was measured in the follow-up period. Second, to account for differences in health outcomes due to differences in reasons for visit during follow-up period, we adjusted for number and types of clinical encounters between measurements of the cardiometabolic risk factors as a time-varying covariate. Third, to account for differences in timing of follow-up visits, we adjusted for COVID-19 transmission in county of residence on dates when cardiometabolic risk factors were measured. COVID-19 cases and test positivity per 100,000 persons in the 7 days prior to the date of measurement were obtained from Centers for Disease Control and Prevention’s COVID-19 County Level of Community Transmission data archive. New cases and test positivity would be zero on all dates for the Historical Control cohort. Finally, we used multiple imputation (5 datasets) to impute missing values of laboratory parameters in lookback period and adjusted for imbalanced covariates in the regression model.

All analysis was carried out using R 4.2.0 and Python 3.11.4.

## Results

The study sample consisted of 249,743 adults, of whom 44,436 (19.2%) belonged to the Exposed cohort, 164,118 (61.5%) belonged to the Unexposed cohort, and 41,189 (19.3%) belonged to the Historical cohort (**Table 1**). Most of the study sample were women (155,223 [62%]), with an average age of 51.9 years (SD: 18.9). Nearly half the sample belonged to minority race-ethnicity groups (Non-Hispanic Black: 21.5%, Hispanic: 21.4%, Non-Hispanic Other: 5.6%). Most patients used private insurance (46%) or Medicare (18%) on index date and 27% were hospitalized on index date. The average BMI on index date in the study sample was 29.5 kg/m^2^ (SD: 7.4) with 30% and 40% having overweight and obesity respectively. The excluded sample without weight available in the preceding year were healthier than the study sample with weight available (**Supplementary Table 2**).

**Table 1.** Socio-demographic, clinical and community characteristics of exposure cohorts before weighting and population standardized bias after weighting.

|  | **Overall**<br>**(n = 249,743)** | **Unweighted** |  |  | **Population**<br>**Standardized**<br>**Bias** |
| --- | --- | --- | --- | --- | --- |
|  |  | **Exposed**<br>**(n = 44,436)** | **Unexposed**<br>**(n = 164,118)** | **Historical**<br>**(n = 41,189)** |  |
| Female (%) | 155,223 (62%) | 29,028 (65%) | 99,949 (61%) | 26,246 (64%) | 0.051 |
| Age (years) | 51.9 (18.9) | 48.8 (18.4) | 53.6 (19.0) | 48.7 (18.4) | **0.186** |
| **Race/ethnicity** |  |  |  |  |  |
| NH White | 128,613 (51.5%) | 19,554 (44.0%) | 88,129 (53.7%) | 20,930 (50.8%) |  |
| NH Black | 53,702 (21.5%) | 11,919 (26.8%) | 32,591 (19.9%) | 9,192 (22.3%) | **0.108** |
| Hispanic | 53,354 (21.4%) | 10,581 (23.8%) | 34,038 (20.7%) | 8,735 (21.2%) | 0.059 |
| NH Other | 14,023 (5.6%) | 2,352 (5.3%) | 9,340 (5.7%) | 2,331 (5.7%) | 0.01 |
| **Smoking** | 117,593 (47%) | 18,818 (42%) | 78,470 (48%) | 20,305 (49%) | **0.162** |
| **Health System** |  |  |  |  |  |
| Source 1 | 92,403 (42%) | 15,492 (36%) | 61,533 (46%) | 15,378 (37%) | **0.127** |
| Source 2 | 51,650 (24%) | 8,225 (19%) | 32,151 (24%) | 11,274 (27%) | **0.168** |
| Source 3 | 24,524 (11%) | 5,429 (12%) | 15,432 (12%) | 3,663 (8.9%) | 0.02 |
| Source 4 | 19,041 (8.7%) | 6,148 (14%) | 8,960 (6.7%) | 3,933 (9.5%) | 0.057 |
| Source 5 | 12,263 (5.6%) | 2,811 (6.5%) | 4,718 (3.6%) | 4,734 (11%) | 0.062 |
| Source 6 | 988 (0.5%) | 187 (0.4%) | 539 (0.4%) | 262 (0.6%) | 0.01 |
| Source 7 | 16,775 (7.7%) | 5,265 (12%) | 9,565 (7.2%) | 1,945 (4.7%) | 0.087 |
| Missing | 32,099 | 879 | 31,220 | 0 |  |
| **Primary Insurance** |  |  |  |  |  |
| Medicare | 38,768 (18%) | 5,809 (13%) | 25,135 (19%) | 7,824 (19%) | 0.014 |
| Medicaid | 18,154 (8.3%) | 3,365 (7.7%) | 11,196 (8.4%) | 3,593 (8.7%) | 0.007 |
| Other Government | 5,299 (2.4%) | 1,257 (2.9%) | 3,314 (2.5%) | 728 (1.8%) | 0.016 |
| Private | 99,648 (46%) | 23,248 (53%) | 54,879 (41%) | 21,521 (52%) | 0.02 |
| No Insurance | 4,210 (1.9%) | 568 (1.3%) | 1,895 (1.4%) | 1,747 (4.2%) | 0.022 |
| No Information | 51,565 (24%) | 9,310 (21%) | 36,479 (27%) | 5,776 (14%) | 0.025 |
| Missing | 32,099 | 879 | 31,220 | 0 |  |
| **Hospitalization within**<br>**30 days of index date** | 66,386 (27%) | 12,506 (28%) | 50,830 (31%) | 3,050 (7.4%) | **0.36** |
| **Diagnosis codes for comorbidities within last 2 years** |  |  |  |  |  |
| Obesity | 10,234 (4.1%) | 2,277 (5.1%) | 6,060 (3.7%) | 1,897 (4.6%) | 0.038 |
| Cardiovascular | 12,533 (5.0%) | 2,179 (4.9%) | 7,915 (4.8%) | 2,439 (5.9%) | 0.019 |
| Cerebrovascular | 549 (0.2%) | 106 (0.2%) | 306 (0.2%) | 137 (0.3%) | 0.007 |
| Hypertension | 23,882 (9.6%) | 4,259 (9.6%) | 14,545 (8.9%) | 5,078 (12%) | 0.027 |
| Pulmonary disease | 7,518 (3.0%) | 1,520 (3.4%) | 4,771 (2.9%) | 1,227 (3.0%) | 0.038 |
| Hyperlipidemia | 18,607 (7.5%) | 3,669 (8.3%) | 11,108 (6.8%) | 3,830 (9.3%) | 0.025 |
| **Medication (any within last 1 year)** |  |  |  |  |  |
| Antidepressants | 32,214 (13%) | 5,988 (13%) | 21,003 (13%) | 5,223 (13%) | 0.027 |
| Antipsychotics | 11,011 (4.4%) | 2,240 (5.0%) | 7,199 (4.4%) | 1,572 (3.8%) | 0.035 |
| Antihypertensives | 68,912 (28%) | 12,560 (28%) | 46,792 (29%) | 9,560 (23%) | 0.093 |
| Statins | 32,296 (13%) | 5,776 (13%) | 22,422 (14%) | 4,098 (9.9%) | 0.098 |
| Immunosuppressants | 22,481 (9.0%) | 4,762 (11%) | 14,734 (9.0%) | 2,985 (7.2%) | 0.049 |
| **Anthropometry within last 1 year** |  |  |  |  |  |
| Height | 66.2 (4.0) | 66.1 (4.0) | 66.2 (4.1) | 66.1 (4.0) | 0.015 |
| *Missing* | 1,442 | 362 | 971 | 109 |  |
| BMI | 29.5 (7.4) | 30.5 (7.7) | 29.3 (7.3) | 29.0 (7.2) | **0.117** |
| Normal (18.5-24.9 kg/m<sup>2</sup>) | 67,955 (27%) | 10,467 (24%) | 45,316 (28%) | 12,172 (30%) |  |
| Overweight (25.0-29.9 kg/m<sup>2</sup>) | 75,092 (30%) | 12,516 (28%) | 50,059 (31%) | 12,517 (30%) |  |
| Obesity (≥ 30 kg/m<sup>2</sup>) | 100,560 (40%) | 20,572 (46%) | 64,419 (39%) | 15,569 (38%) |  |
| Systolic BP | 125.8 (18.6) | 125.1 (17.9) | 126.0 (19.0) | 125.7 (17.9) | 0.017 |
| *Missing* | 30,502 | 7,846 | 19,530 | 3,126 |  |
| **Labs within last 1 year** |  |  |  |  |  |
| HbA1c (%) | 6.3 (1.7) | 6.4 (1.8) | 6.2 (1.6) | 6.2 (1.6) | **0.141** |
| *Missing* | 201,296 | 33,909 | 131,239 | 36,148 |  |
| Glucose (mg/dL) | 113.6 (47.3) | 114.7 (49.7) | 113.9 (47.2) | 110.4 (43.4) | 0.094 |
| *Missing* | 108,989 | 17,165 | 68,056 | 23,768 |  |
| ALT | 17.0 (12.0, 26.0) | 17.0 (12.0, 26.0) | 17.0 (12.0, 26.0) | 17.0 (12.0, 26.0) | 0.032 |
| *Missing* | 130,240 | 20,512 | 82,511 | 27,217 |  |
| AST | 20.0 (16.0, 26.0) | 19.0 (15.0, 25.0) | 20.0 (16.0, 26.0) | 19.0 (16.0, 25.0) | 0.057 |
| *Missing* | 128,790 | 20,166 | 81,843 | 26,781 |  |
| Serum Creatinine | 0.8 (0.7, 1.1) | 0.8 (0.7, 1.0) | 0.9 (0.7, 1.1) | 0.8 (0.7, 1.0) | **0.109** |
| *Missing* | 111,019 | 17,398 | 69,416 | 24,205 |  |
| HDL | 50.8 (16.9) | 50.1 (15.8) | 50.9 (16.8) | 51.7 (19.2) | **0.103** |
| *Missing* | 194,042 | 32,716 | 127,000 | 34,326 |  |
| LDL | 83.1 (49.9) | 80.4 (53.5) | 81.6 (49.7) | 95.9 (42.0) | **0.393** |
| *Missing* | 191,750 | 31,939 | 125,748 | 34,063 |  |

Medication and Comorbidities were imputed as ‘No’ if data were not found in PRESCRIBING and DIAGNOSIS datasets.
Hospitalization within 30 days of index date was ‘Yes’ if any encounter of type emergency department to inpatient (ED), inpatient (IP), non-acute institutional stay (IS) and observation stay (OS). All others were coded as ‘No’.
Values were Mean (standard deviation) or median (25<sup>th</sup> percentile, 75<sup>th</sup> percentile) or frequency (percentage%)

The Exposed cohort was younger (48.8 years [SD: 18.4]) and more likely to be female (65%) relative to the Unexposed cohort (age: 53.6 years [SD: 19.0], 61% female). The Exposed cohort was also more likely to be Non-Hispanic Black or Hispanic race-ethnicity (50.6% vs 40.6% Unexposed and 43.5% Historical), privately insured (53% vs 42% Unexposed and 42% Historical) and had higher obesity (46% vs 39% Unexposed and 38% Historical). The Unexposed cohort was more likely to be hospitalized due to any cause relative to other cohorts (31% vs 28% Exposed and 7.4% Historical). The proportion with comorbidities and medications prescribed were similar between the three cohorts. After inverse probability weighting (**Supplementary Table 3**), some covariates remained imbalanced between the three groups, namely age, race-ethnicity, smoking, health system at admission, hospitalization status, BMI, and laboratory parameters (HbA1c, serum creatinine, HDL, LDL) in the previous year.

### Trajectories of body mass index in follow-up period

Of the study sample, only 194,792 (77.9%) had BMI information in the follow-up period (Exposed: 32,384, Unexposed: 127,142, Historical: 35,266). Socio-demographic and clinical characteristics were similar between those with and without BMI information in the follow-up period (**Supplementary Table 2**). The random forests model to predict loss to follow-up had high model fit in the hold-out test set (AUROC: 0.97; **Supplementary Methods Note**). The time (median [IQR]) to the first follow-up visit of BMI was shorter for Exposed (39 [12, 107]) and Unexposed (33 [11, 98]) cohorts, relative to the Historical (74 [22, 180]) cohort (**Supplementary Table 4**). The Historical cohort had more frequent (median: 4 vs 3 each for Exposed and Unexposed) and longer duration of follow-up (median days [IQR]; 464 [271, 592] vs 178 [76, 357] for Exposed and 212 [85, 374] for Unexposed). BMI at the first follow-up was higher for the Exposed (29.3 kg/m^2^, 95% Confidence Interval [CI]: 24.9, 34.9), relative to Unexposed (27.9 kg/m^2^, 95%CI: 23.9, 33.0) and Historical (27.8 kg/m^2^, 95%CI: 23.8, 32.8) cohorts.

Adjusting for imbalanced covariates and after IPW, the average estimated BMI at origin date (**Table 2**) was higher for the Exposed (29.3 kg/m^2^, 95%CI: 29.0, 29.7) relative to the Unexposed (difference: –0.07 kg/m^2^, 95%CI: –0.12, –0.01) and Historical (difference: –0.27 kg/m^2^, 95%CI: –0.34, –0.20) cohorts. Change in BMI per 100 days of follow-up was null for the Exposed (0.00 kg/m^2^, 95%CI: –0.03, 0.03), negative for the Unexposed (–0.03 kg/m^2^, 95%CI: – 0.04, –0.02) and positive for the Historical (0.04 kg/m^2^, 95%CI: 0.03, 0.05) cohort. The change in BMI per 100 days of follow-up for the Unexposed (–0.04 kg/m^2^, 95%CI: –0.06, –0.01) and Historical (0.03 kg/m^2^, 95%CI: 0.00, 0.06) cohorts were different, relative to the Exposed cohort (**Table 2**).

**Table 2.** Marginal estimates at origin date and change at 100 days of follow-up for body mass index.

|  | **Exposed** | **Unexposed** | **Historical** |
| --- | --- | --- | --- |
| $\text{Time}_{0,\text{Cohort}} - \text{Time}_{0,\text{Exposed}}$ | Ref | -0.07 (-0.12, -0.01) | -0.27 (-0.34, -0.20) |
| $\Delta_{0 \rightarrow 100\text{days}, \text{Cohort}}$ | 0 (-0.03, 0.03) | -0.04 (-0.05, -0.02) | 0.03 (0.01, 0.04) |
| $\Delta_{0 \rightarrow 100\text{days}, \text{Cohort}} - \Delta_{0 \rightarrow 100\text{days}, \text{Exposed}}$ | Ref | -0.04 (-0.06, -0.01) | 0.03 (0, 0.06) |

All estimates are from the marginal structural model with statistical interaction of exposure group, socio-demographic characteristic (i.e., sex, age, race-ethnicity) and time, after inverse probability weighting for confounding and loss to follow-up. Associations are adjusted for BMI within last 1 year of lookback period and imbalanced covariates.

Adjusted differences in BMI at origin date for subgroups of sex, age, race-ethnicity, and hospitalization status are presented in **Figure 1A**. Across most socio-demographic subgroups (except among adult Males, those aged 65 and older, Hispanic adults, non-hospitalized adults) BMI at origin date was higher among Exposed, relative to Unexposed and Historical cohorts (**Supplementary Table 5**). Relative to Exposed cohort (**Figure 1B**), the change in BMI per 100 days of follow-up among the Unexposed cohort was lower among Males (difference-in-differences [DiD]: –0.07 kg/m^2^, 95%CI: –0.12, –0.02) and non-Hospitalized adults (DiD: –0.04 kg/m^2^, 95%CI: –0.07, –0.01). Relative to the Exposed cohort, the change in BMI per 100 days of follow-up among the Historical cohort was higher among Females (DiD: 0.07 kg/m^2^, 95%CI: 0.03, 0.11), Non-Hispanic White adults (DiD: 0.07 kg/m^2^, 95%CI: 0.03, 0.11) and non-Hospitalized adults (DiD: 0.03 kg/m^2^, 95%CI: 0.00, 0.06)

**Figure 1.**
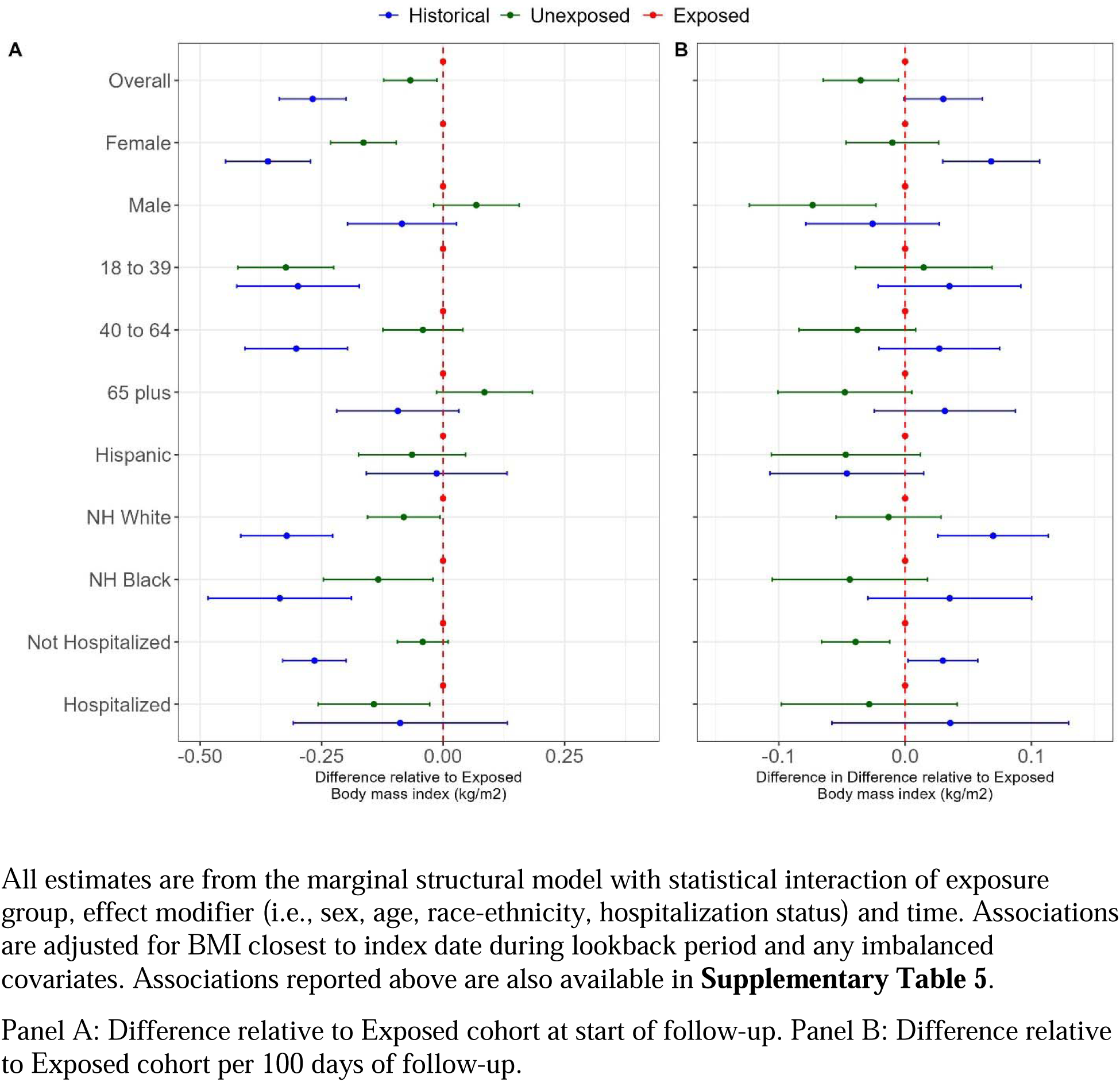
Difference from Exposed cohort in body mass index at start of post-acute period and change per 100 days of follow-up for Unexposed and Historical cohorts.

Results were similar when restricting to those with at least 100 days of follow-up and at least two encounters when BMI was measured (**Supplementary Table 6**), after adjusting for number and types of encounters between BMI measurements (**Supplementary Table 7**) and after adjusting for COVID-19 cases and testing (**Supplementary Table 8**). Results were also similar after accounting for missing data in laboratory parameters using multiple imputation **(Supplementary Table 9**).

## Discussion

In a diverse real-world cohort of adults who sought medical care during the pandemic, those who tested positive for COVID-19 (Exposed) were heavier at the start of the post-acute phase of SARS-CoV-2 infection compared to contemporary controls who tested negative (Unexposed), and age-sex matched historical controls. The BMI of those Exposed did not change per 100 days of follow-up, while the BMI of those Unexposed decreased, and the BMI of historical controls increased. The change in BMI over time differed between Exposed and Unexposed cohorts at start of follow-up among Males and those 65 years or older, and in changes per 100 days among Males and Hispanics.

The persistent higher BMI, including before infection, among those Exposed compared to those who were Unexposed suggest that those infected by SARS-CoV-2 may have been at higher risk of COVID-19 when seeking care.^5^ This finding is consistent with numerous reports of elevated BMI being a risk factor for severe COVID-19.^24,25^ The current study extends the findings of this prior research to examine BMI trajectories following SARS-CoV-2 infection at the individual level. To date, most studies have examined the pandemic’s impact on BMI at a population level. For instance, a meta-analysis of observational studies showed that the pandemic resulted in a small increase in BMI and prevalence of obesity among adults.^26,27^ Potential explanations for BMI increases at the individual and population level include rise in sedentary time and unhealthy dietary habits, downward social mobility from loss of income and employment, and disruption of preventive care.^26^ The bidirectional relationship of viral infections and cardiometabolic disease may be mediated by excess adiposity, insulin resistance, inflammation and/or delayed recovery after COVID-19 as a post-acute sequelae.^2–4,28,29^.

This analysis of real-world data has several strengths. We used a validated computable phenotype for prevalent T2DM as part of our exclusion criteria to identify a cohort of adults free of T2DM, who were users of the healthcare system before the pandemic, and who were tested for SARS-CoV-2. We additionally included a historical control cohort, matched on age and sex, to characterize differences in the patient population attributable to the pandemic. The OneFlorida+ Data Trust is a part of the National Patient-Centered Clinical Research Network (PCORnet) and follows the common data model, facilitating reproducibility. Compared to other real-world studies, the analytic sample was younger and more diverse across categories of race-ethnicity, age, and sex.^2,5^ The duration of follow-up was longer than other studies reporting post-acute sequelae of COVID-19.^3,5,6,13^ We used methods for target trial emulation to minimize confounding, including minimizing the potential for instrumental variable bias in high dimensional confounder selection.

There are several limitations for this study. First, the OneFlorida+ CRN includes data from many but not all of Florida’s healthcare providers; our analyses could not account for patients testing positive at centers not participating in the OneFlorida+ CRN. However, the criteria we used for the lookback period were meant to identify regular users of healthcare systems participating in OneFlorida+.^30^ Additionally, our sample selection was not biased by home testing because it became widespread only after the last date of follow-up in February 2022. Second, although we used a high dimensional propensity score for confounding adjustment, we cannot rule out all differences in clinical characteristics at the time of the index encounter. For instance, those Exposed and Unexposed may have had different reasons for seeking care. Finally, singly-robust machine learning based propensity scores are inferior to doubly-robust methods for lower bias and confidence-interval coverage.^31^ However, to our knowledge, there were no standard software packages to implement doubly-robust causal inference methods for sparse, longitudinal datasets such as electronic health records.

This analysis of diverse, real-world data showed that among those who sought medical care during the pandemic, patients testing positive were heavier at the start of the post-acute phase of COVID-19 and retained weight during the post-acute phase of follow-up compared to those testing negative throughout the observation period. Although the subgroup analysis revealed some heterogeneity in these associations, the observed associations were present among racial-ethnic minorities and young adults who were disproportionately affected by COVID-19. The mechanisms influencing this observed increase in weight retention after COVID-19 infection warrant further investigation.

## Supporting information

RECORD-PE Checklist

## Abbreviations

BMI: Body mass index
CP: Computable phenotype
EHR: Electronic health record
HDL: High density lipoprotein
LDL: Low density lipoprotein
PASC: Post-acute sequelae of COVID-19
T2DM: Type 2 diabetes

## ACKNOWLEDGEMENT/SUPPORT

Acknowledgements

This research was supported by the National Institute for Diabetes and Digestive and Kidney Diseases (NIDDK) of the National Institutes of Health, award number 3R01DK120814-05S1. MKA was partially supported by the Georgia Center for Diabetes Translation Research which is funded by the National Institutes of Health (P30DK111024). The authors thank the OneFlorida+ Data Trust team (Kathryn Shaw, Meggen Kaufman, Jiang Bian, Elizabeth Shenkman) for support on query development and data extraction. The authors thank Shihab Chowdhury for administrative support.

## Author contributions

RJC, WTD and JSV conceptualized the study with inputs from MKA and YG. JSV conducted the analysis. JSV wrote the first draft with inputs from RJC. All authors reviewed and edited subsequent drafts.

## Data availability statement

The code for the analysis is available on https://github.com/jvargh7/pasc_cardiometabolic_risk. Information of the OneFlorida+ CRN is provided at https://onefloridaconsortium.org/, and OneFlorida+ data are made available to researchers with an approved study protocol at https://onefloridaconsortium.org/front-door/prep-to-research-data-query/. For questions regarding OneFlorida+.

## Figure 1 Legend

Our historical cohort corresponds with the blue lines. Our unexposed cohort corresponds with the green lines. Our exposed cohort corresponds with the red lines.

