## Supplementary Material for "Body mass index changes and their association with SARS-CoV-2 infection: a real-world analysis"

**Supplementary Table 1. Definitions of comorbidities and medication classes during look-back and follow-up period**

| | Codes included |
|---|---|
| **Exposure Definition** | |
| COVID-19 Lab | 94500-6,94309-2,94558-4,94534-5,94759-8,95209-3,95608-6,95380-2,94316-7,95422-2,94533-7,94565-9,94756-4,94642-6,94306-8,95406-5,95425-5,87635 |
| COVID-19 ICD10-CM | U07.1, U07.2, J12.81, J12.82, B34.2, B97.2, B97.21, B97.29, U04, U04.9 |
| **Data for Diabetes Diagnosis (Weise 2018)** | |
| HbA1c | 4548-4, 41995-2, 55454-3, 71875-9, 549-2, 17856-6, 59261-6, 62388-4, 17855-8 |
| Diabetes | E11.00, E11.01, E11.21, E11.22, E11.29, E11.311, E11.319, E11.321, E11.329, E11.331, E11.339, E11.341, E11.349, E11.351, E11.359, E11.36, E11.39, E11.40, E11.41, E11.42, E11.43, E11.44, E11.49, E11.51, E11.52, E11.59, E11.610, E11.618, E11.620, E11.621, E11.622, E11.628, E11.630, E11.638, E11.641, E11.649, E11.65, E11.69, E11.8, E11.9 |
| Sulfonylurea | tolazamide [Tolinase]; acetohexamide [Dymelor]; tolbutamide [Orinase, Orinase Diagnostic, Tol-Tab], chlorpropamide [Diabinese] [diabanase][diabinase]; glipizide [Glucotrol, Glucotrol XL, Glucatrol]; glyburide [Micronase, Glynase, Diabetamide, Diabeta]; glimepiride [Amaryl] |
| Biguanide | metformin [Glucophage] |
| Meglitanide | repaglinide [Prandin]; nateglinide [Starlix] |
| Alpha glucosidase Inhibitor | miglitol [Glyset]; acarbose [Precose] |
| Thiazolidinedione | rosiglitazone [Avandia]; pioglitazone [ACTOS]; troglitazone [Rezulin] |
| Sodium-glucose cotransporter-2 Inhibitor | dapagliflozin [Farxiga]; canagliflozin [Invokana or Sulisent]; empagliflozin [Jardiance], ertugliflozin [Segluromet or Steglatro or Steglujan] |
| Dipeptidyl peptidase-4 Inhibitors | sitagliptin [Januvia]; saxagliptin [Onglyza]; linagliptin [Tradjenta], alogliptin [Kazano or Nesina or Oseni] |
| Glucose-dependent insulintropic peptide-1 | exenatide [Byetta]; liraglutide [Victoza], lixisenatide [Adlyxin or Soliqua], semaglutide [Rybelsus or Ozempic or Wegovy] |

| | |
|---|---|
| Insulin | regular insulin [Humulin R, Novolin R]; insulin lispro [Humalog]; insulin aspart [Novolog]; insulin glulisine [Apidra]; prompt insulin zinc [Semilente]; isophane insulin, neutral protamine hagedorn [NPH] [Humulin N, Novolin N]; insulin zinc [Lente]; extended insulin zinc insulin [Ultralente]; insulin glargine [Lantus]; insulin detemir [Levemir] |
| **Comorbidities (Bachmann 2020)** | |
| Cerebrovascular – Transient Ischemic Attack | G45.0; G45.1; G45.8; G45.9; I67.848 |
| Cerebrovascular – Stroke | I67.89; I60.9; I61.9; I63.30; I63.40; I63.50; I66.09; I66.19; I66.29; I66.9; I67.89 |
| Cardiovascular – Atrial Fibrillation | I48.91; I48.92 |
| Cardiovascular – Arrhythmia | I44.*; I45.*; I47.*; I49.* |
| Cardiovascular – Coronary Artery Disease | I24.*; I25.*; I20.* |
| Cardiovascular – Congestive Heart Failure | I11.0; I13.0; I13.2; I50.9; I50.1; I50.20; I50.21; I50.22; I50.23; I50.30; I50.31; I50.32; I50.33; I50.40; I50.41; I50.42; I50.43 |
| Cardiovascular – Cardiac valve disease | I05.*; I06.*; I08.*; I34.*; I35.* |
| Cardiovascular – Peripheral artery disease, revascularization, peripheral vasodilators | I70.2*; I72.*; I77.*; I73.9; I75.* |
| Hypertension | I10; I11.*; I12.*; I13.* |
| Pulmonary – General | J96.*; R092; I26.9*; I27.* |
| Pulmonary – COPD/ Emphysema/Asthma | J41.0; J41.1; J44.9; J44.1; J44.0; J41.8; J42; J43; J43.0; J43.1; J43.2; J43.8; J43.9; J45.20; J45.22; J45.21; J45.990; J45.991; J45.909; J45.998; J45.902; J45.901; Z13.83 |
| Lipid disorders | E78.* |
| Obesity | E66.0; E66.1; E66.2; E66.8; E66.9 |
| **Labs** | |
| LDL | 13457-7, 18262-6, 2089-1, 11054-4 |
| HDL | 18263-4, 2085-9 |
| Triglycerides | 12951-0, 2571-8 |
| ALT | 1742-6, 1744-2 |
| AST | 1920-8 |

| | |
|---|---|
| HbA1c | 4548-4, 41995-2, 55454-3, 71875-9, 549-2, 17856-6, 59261-6, 62388-4, 17855-8 |
| Serum Creatinine | 2160-0 |
| Glucose | 2345-7, 2339-0, 41653-7, 2340-8, 27353-2, 1547-9, 1558-6 |
| **Encounters** | Types of encounters included clinical (ambulatory, inpatient, observation stay, emergency department, emergency to inpatient, institutional professional consult, non-acute institutional stay, telehealth), laboratory (other ambulatory, unknown, no information, and other encounters and matched to lab result) and pharmacy (other ambulatory, unknown, no information, and other encounters matched to prescription data) visits. |

**Supplementary Table 2. Socio-demographic characteristics by availability of follow-up information (n = 388,060)**

| | | | | Lookback available | |
|---|---|---|---|---|---|
| | **Overall** | **Lookback available** | **Lookback not available** | **Available during both lookback and follow-up** | **Only lookback** |
| *N* | 388,060 | 249,743 | 138,317 | 194,792 | 54,951 |
| **Exposure Group** | | | | | |
| Exposed | 74,629 (19%) | 44,436 (18%) | 30,193 (22%) | 32,384 (17%) | 12,052 (22%) |
| Unexposed | 238,499 (61%) | 164,118 (66%) | 74,381 (54%) | 127,142 (65%) | 36,976 (67%) |
| Historical | 74,932 (19%) | 41,189 (16%) | 33,743 (24%) | 35,266 (18%) | 5,923 (11%) |
| Female (%) | 241,787 (62%) | 155,223 (62%) | 86,564 (63%) | 120,709 (62%) | 34,514 (63%) |
| Age (years) | 51.0 (19.2) | 51.9 (18.9) | 49.2 (19.6) | 52.1 (18.6) | 51.4 (19.8) |
| **Race/ethnicity** | | | | | |
| NH White | 195,734 (50.4%) | 128,613 (51.5%) | 67,121 (48.5%) | 101,352 (52.0%) | 27,261 (49.6%) |
| NH Black | 78,115 (20.1%) | 53,702 (21.5%) | 24,413 (17.7%) | 43,169 (22.2%) | 10,533 (19.2%) |
| Hispanic | 87,189 (22.5%) | 53,354 (21.4%) | 33,835 (24.5%) | 40,025 (20.5%) | 13,329 (24.3%) |
| NH Other | 25,617 (6.6%) | 14,023 (5.6%) | 11,594 (8.4%) | 10,221 (5.2%) | 3,802 (6.9%) |
| **Smoking** | | | | | |
| Yes | 124,816 (44%) | 117,593 (47%) | 7,223 (23%) | 103,185 (53%) | 14,408 (26%) |
| Missing | 106,447 | 0 | 106,447 | | |
| **Health System** | | | | | |
| Source 1 | 105,379 (31%) | 92,403 (42%) | 12,976 (11%) | 84,682 (49%) | 7,721 (17%) |
| Source 2 | 77,730 (23%) | 51,650 (24%) | 26,080 (21%) | 39,459 (23%) | 12,191 (26%) |
| Source 3 | 47,066 (14%) | 24,524 (11%) | 22,542 (19%) | 14,521 (8.5%) | 10,003 (22%) |
| Source 4 | 41,752 (12%) | 19,041 (8.7%) | 22,711 (19%) | 12,780 (7.5%) | 6,261 (14%) |
| Source 5 | 15,605 (4.6%) | 12,263 (5.6%) | 3,342 (2.7%) | 9,442 (5.5%) | 2,821 (6.1%) |
| Source 6 | 1,428 (0.4%) | 988 (0.5%) | 440 (0.4%) | 794 (0.5%) | 194 (0.4%) |
| Source 7 | 50,483 (15%) | 16,775 (7.7%) | 33,708 (28%) | 9,722 (5.7%) | 7,053 (15%) |
| Missing | 48,617 | 32,099 | 16,518 | 23,392 | 8,707 |
| **Primary Insurance** | | | | | |
| Medicare | 52,058 (15%) | 38,768 (18%) | 13,290 (11%) | 32,680 (19%) | 6,088 (13%) |
| Medicaid | 23,333 (6.9%) | 18,154 (8.3%) | 5,179 (4.3%) | 15,901 (9.3%) | 2,253 (4.9%) |
| Other Government | 7,838 (2.3%) | 5,299 (2.4%) | 2,539 (2.1%) | 4,417 (2.6%) | 882 (1.9%) |
| Private | | 99,648 (46%) | | 78,890 (46%) | 20,758 (45%) |
| No Insurance | 7,408 (2.2%) | 4,210 (1.9%) | 3,198 (2.6%) | 3,248 (1.9%) | 962 (2.1%) |
| No Information | 98,498 (29%) | 51,565 (24%) | 46,933 (39%) | 36,264 (21%) | 15,301 (33%) |
| Missing | 48,617 | 32,099 | 16,518 | 23,392 | 8,707 |
| **Hospitalization within 30 days of index date** | 99,063 (26%) | 66,386 (27%) | 32,677 (24%) | 49,380 (25%) | 17,006 (31%) |
| **Medication (any within last 1 year)** | | | | | |
| Antidepressants | 38,114 (9.8%) | 32,214 (13%) | 5,900 (4.3%) | 27,737 (14%) | 4,477 (8.1%) |
| Antipsychotics | 12,059 (3.1%) | 11,011 (4.4%) | 1,048 (0.8%) | 9,547 (4.9%) | 1,464 (2.7%) |
| Antihypertensives | 80,737 (21%) | 68,912 (28%) | 11,825 (8.5%) | 57,025 (29%) | 11,887 (22%) |
| Statins | 37,744 (9.7%) | 32,296 (13%) | 5,448 (3.9%) | 26,857 (14%) | 5,439 (9.9%) |

| | | | | | |
|---|---|---|---|---|---|
| Immunosuppressants | 26,771 (6.9%) | 22,481 (9.0%) | 4,290 (3.1%) | 17,269 (8.9%) | 5,212 (9.5%) |
| **Anthropometry closest within last 1 year** | | | | | |
| Height | 66.2 (4.0) | 66.2 (4.0) | NA (NA) | 66.2 (4.0) | 66.1 (4.0) |
| *Missing* | 139,759 | 1,442 | 138,317 | 0 | 1,442 |
| BMI | 29.5 (7.4) | 29.5 (7.4) | NA (NA) | 29.5 (7.4) | 29.4 (7.4) |
| *Missing* | 138,317 | 0 | 138,317 | | |
| Systolic BP | 125.9 (18.6) | 125.8 (18.6) | 126.6 (18.3) | 125.9 (18.6) | 125.3 (18.7) |
| *Missing* | 143,751 | 30,502 | 113,249 | 16,973 | 13,529 |
| **Labs within last 1 year** | | | | | |
| HbA1c (%) | 6.2 (1.6) | 6.3 (1.7) | 6.0 (1.3) | 6.3 (1.7) | 6.2 (1.6) |
| *Missing* | 323,579 | 201,296 | 122,283 | 155,661 | 45,635 |
| Glucose (mg/dL) | 112.8 (46.9) | 113.6 (47.3) | 110.2 (45.8) | 113.8 (47.7) | 113.2 (45.6) |
| *Missing* | 204,147 | 108,989 | 95,158 | 84,112 | 24,877 |
| ALT | 17.0 (12.0, 26.0) | 17.0 (12.0, 26.0) | 18.0 (13.0, 26.0) | 17.0 (12.0, 26.0) | 18.0 (12.0, 27.0) |
| *Missing* | 230,313 | 130,240 | 100,073 | 101,189 | 29,051 |
| AST | 20.0 (16.0, 26.0) | 20.0 (16.0, 26.0) | 20.0 (16.0, 25.0) | 20.0 (16.0, 26.0) | 20.0 (16.0, 26.0) |
| *Missing* | 228,832 | 128,790 | 100,042 | 99,874 | 28,916 |
| Serum Creatinine | 0.8 (0.7, 1.0) | 0.8 (0.7, 1.1) | 0.8 (0.7, 1.0) | 0.9 (0.7, 1.1) | 0.8 (0.7, 1.0) |
| *Missing* | 206,250 | 111,019 | 95,231 | 86,029 | 24,990 |
| HDL | 51.7 (16.7) | 50.8 (16.9) | 54.0 (16.1) | 50.9 (17.0) | 50.6 (16.4) |
| *Missing* | 312,407 | 194,042 | 118,365 | 148,261 | 45,781 |
| LDL | 79.6 (52.0) | 83.1 (49.9) | 70.3 (56.1) | 87.1 (48.2) | 65.3 (53.7) |
| *Missing* | 308,329 | 191,750 | 116,579 | 147,313 | 44,437 |
| **Diagnosis codes for comorbidities within last 2 years** | | | | | |
| Obesity | 12,014 (3.1%) | 10,234 (4.1%) | 1,780 (1.3%) | 8,656 (4.4%) | 1,578 (2.9%) |
| Cardiovascular | 14,797 (3.8%) | 12,533 (5.0%) | 2,264 (1.6%) | 10,325 (5.3%) | 2,208 (4.0%) |
| Cerebrovascular | 684 (0.2%) | 549 (0.2%) | 135 (<0.1%) | 448 (0.2%) | 101 (0.2%) |
| Hypertension | 29,399 (7.6%) | 23,882 (9.6%) | 5,517 (4.0%) | 19,531 (10%) | 4,351 (7.9%) |
| Pulmonary disease | 9,158 (2.4%) | 7,518 (3.0%) | 1,640 (1.2%) | 6,317 (3.2%) | 1,201 (2.2%) |
| Hyperlipidemia | 21,787 (5.6%) | 18,607 (7.5%) | 3,180 (2.3%) | 15,493 (8.0%) | 3,114 (5.7%) |

**Supplementary Table 3. Socio-demographic, clinical and community characteristics of exposure cohorts after weighting**

| | **Overall** | **Exposed** | **Historical** | **Unexposed** |
|---|---|---|---|---|
| *Weighted N* | 190,028 | 25,526 | 149,031 | 15,471 |
| Female (%) | 117,941 (62%) | 16,464 (64%) | 91,688 (62%) | 9,789 (63%) |
| Age (years) | 51.8 (19.0) | 49.4 (18.6) | 52.6 (19.0) | 48.5 (18.4) |
| **Race/ethnicity** | | | | |
| NH White | 98,384 (51.8%) | 11,726 (45.9%) | 78,626 (52.8%) | 8,031 (51.9%) |
| NH Black | 41,248 (21.7%) | 6,639 (26.0%) | 30,970 (20.8%) | 3,639 (23.5%) |
| Hispanic | 39,543 (20.8%) | 5,778 (22.6%) | 30,832 (20.7%) | 2,932 (19.0%) |
| **Smoking** | | | | |
| Yes | 91,930 (48%) | 11,608 (45%) | 71,788 (48%) | 8,534 (55%) |
| **Health System** | | | | |
| Source 1 | 72,287 (44%) | 9,675 (39%) | 55,894 (45%) | 6,718 (43%) |
| Source 2 | 35,373 (22%) | 4,677 (19%) | 27,634 (22%) | 3,063 (20%) |
| Source 3 | 18,667 (11%) | 3,010 (12%) | 14,350 (12%) | 1,308 (8.5%) |
| Source 4 | 14,571 (8.9%) | 3,187 (13%) | 9,649 (7.8%) | 1,734 (11%) |
| Source 5 | 8,945 (5.5%) | 1,619 (6.5%) | 5,797 (4.7%) | 1,529 (9.9%) |
| Source 6 | 789 (0.5%) | 117 (0.5%) | 572 (0.5%) | 100 (0.6%) |
| Source 7 | 13,448 (8.2%) | 2,615 (11%) | 9,813 (7.9%) | 1,020 (6.6%) |
| **Primary Insurance** | | | | |
| Medicare | 28,774 (18%) | 3,508 (14%) | 22,389 (18%) | 2,877 (19%) |
| Medicaid | 13,715 (8.4%) | 1,969 (7.9%) | 10,379 (8.4%) | 1,367 (8.8%) |
| Other Government | 4,223 (2.6%) | 727 (2.9%) | 3,191 (2.6%) | 305 (2.0%) |
| Private | 64,524 (45%) | 10,845 (53%) | 47,759 (42%) | 5,920 (51%) |
| No Insurance | 3,053 (1.9%) | 339 (1.4%) | 2,068 (1.7%) | 647 (4.2%) |
| No Information | 39,675 (24%) | 5,090 (20%) | 32,399 (26%) | 2,186 (14%) |
| **Hospitalization within 30 days of index date** | 52,528 (28%) | 7,125 (28%) | 43,728 (29%) | 1,675 (11%) |
| **Medication (any within last 1 year)** | | | | |
| Antidepressants | 28,094 (15%) | 3,792 (15%) | 22,159 (15%) | 2,143 (14%) |
| Antipsychotics | 10,248 (5.4%) | 1,411 (5.5%) | 8,118 (5.4%) | 719 (4.6%) |
| Antihypertensives | 60,532 (32%) | 8,098 (32%) | 48,141 (32%) | 4,293 (28%) |
| Statins | 29,913 (16%) | 3,880 (15%) | 24,132 (16%) | 1,900 (12%) |
| Immunosuppressants | 20,888 (11%) | 3,149 (12%) | 16,259 (11%) | 1,480 (9.6%) |
| **Anthropometry closest within 1 year** | | | | |
| Height | 66.2 (4.0) | 66.2 (4.0) | 66.2 (4.1) | 66.1 (4.0) |
| *Missing* | 1,250 | 227 | 980 | 43 |
| BMI | 29.5 (7.4) | 30.3 (7.7) | 29.4 (7.4) | 29.2 (7.3) |
| Systolic BP | 125.7 (18.6) | 125.4 (18.0) | 125.7 (18.8) | 125.6 (18.1) |
| *Missing* | 21,232 | 3,499 | 16,550 | 1,183 |
| **Labs within last 1 year** | | | | |
| HbA1c (%) | 6.3 (1.7) | 6.3 (1.8) | 6.2 (1.6) | 6.2 (1.6) |
| *Missing* | 152,493 | 19,935 | 119,298 | 13,260 |

| | | | | |
|---|---|---|---|---|
| Glucose (mg/dL) | 113.9 (47.5) | 114.7 (49.2) | 114.0 (47.5) | 110.9 (44.1) |
| *Missing* | 80,265 | 10,435 | 61,998 | 7,832 |
| ALT | 17.0 (12.0, 26.0) | 17.0 (12.0, 26.0) | 17.0 (12.0, 26.0) | 17.0 (12.0, 26.0) |
| *Missing* | 96,773 | 12,490 | 74,991 | 9,292 |
| AST | 20.0 (16.0, 26.0) | 19.0 (15.0, 25.0) | 20.0 (16.0, 26.0) | 19.0 (16.0, 25.0) |
| *Missing* | 95,684 | 12,306 | 74,227 | 9,151 |
| Serum Creatinine | 0.9 (0.7, 1.1) | 0.8 (0.7, 1.0) | 0.9 (0.7, 1.1) | 0.8 (0.7, 1.0) |
| *Missing* | 81,720 | 10,589 | 63,149 | 7,981 |
| HDL | 50.8 (16.8) | 50.3 (15.8) | 50.8 (16.8) | 51.8 (18.9) |
| *Missing* | 146,292 | 19,095 | 114,736 | 12,461 |
| LDL | 83.1 (49.7) | 83.4 (52.1) | 82.0 (49.7) | 94.6 (43.0) |
| *Missing* | 144,722 | 18,760 | 113,586 | 12,376 |
| **Diagnosis codes for comorbidities within last 2 years** | | | | |
| Obesity | 7,409 (3.9%) | 1,180 (4.6%) | 5,705 (3.8%) | 524 (3.4%) |
| Cardiovascular | 9,113 (4.8%) | 1,222 (4.8%) | 7,171 (4.8%) | 719 (4.6%) |
| Cerebrovascular | 376 (0.2%) | 53 (0.2%) | 285 (0.2%) | 38 (0.2%) |
| Hypertension | 17,168 (9.0%) | 2,369 (9.3%) | 13,323 (8.9%) | 1,477 (9.5%) |
| Pulmonary disease | 5,608 (3.0%) | 805 (3.2%) | 4,433 (3.0%) | 371 (2.4%) |
| Hyperlipidemia | 13,253 (7.0%) | 1,990 (7.8%) | 10,181 (6.8%) | 1,083 (7.0%) |

Values are summarized as weighted mean (weighted SD) or weighted median (weighted 25$^{th}$ percentile, weighted 75$^{th}$ percentile) or weighted frequency (weighted %).

**Supplementary Table 4. Changes during follow-up period in body mass index**

| Body mass index | Exposed<br>Mar 2020 – Feb 2022 | Unexposed<br>Mar 2020 – Feb 2022 | Historical<br>Mar 2018 – Feb 2020 |
|---|---|---|---|
| Number of participants with any observation in follow-up | 32,384 | 127,142 | 35,266 |
| Time to first follow-up visit (days relative to origin date [T0 + 30]) | 39 [12, 107] | 33 [11, 98] | 74 [22, 180] |
| BMI at first follow-up encounter after T0+30 days ($kg/m^2$) | 29.3 [24.9, 34.9] | 27.9 [23.9, 33] | 27.8 [23.8, 32.8] |
| Number of BMI measurements after T0+30 | 3 [1, 6] | 3 [1, 7] | 4 [2, 8] |
| Length of follow-up (days relative to origin date [T0 + 30]) | 178 [76, 357] | 212 [85, 374] | 464 [271, 592] |
| Maximum change in BMI during follow-up ($kg/m^2$) | 0.9 [0, 2.6] | 1.1 [0, 2.7] | 1.3 [0.2, 3] |

All values are unweighted median ($25^{th}$ percentile, $75^{th}$ percentile)

**Supplementary Table 5. Modeled marginal estimates of changes in body mass index during follow-up by exposure cohort**

| | | **Exposed** | **Unexposed** | **Historical** |
|---|---|---|---|---|
| Female | $Time_{0,Cohort}$ | 29.8 (29.4, 30.2) | 29.6 (29.4, 29.8) | 29.4 (29, 29.8) |
| | $Time_{0,Cohort}$ - $Time_{0,Exposed}$ | Ref | **-0.16 (-0.23, -0.1)** | **-0.36 (-0.45, -0.27)** |
| | $\Delta_{0\rightarrow 100days}$ | -0.02 (-0.05, 0.01) | -0.03 (-0.05, -0.02) | 0.05 (0.03, 0.07) |
| | $\Delta_{0\rightarrow 100days, Cohort}$ – $\Delta_{0\rightarrow 100days, Exposed}$ | Ref | -0.01 (-0.05, 0.03) | **0.07 (0.03, 0.11)** |
| Male | $Time_{0,Cohort}$ | 29.5 (29, 30.1) | 29.6 (29.4, 29.9) | 29.5 (28.9, 30) |
| | $Time_{0,Cohort}$ - $Time_{0,Exposed}$ | Ref | 0.07 (-0.02, 0.16) | -0.08 (-0.2, 0.03) |
| | $\Delta_{0\rightarrow 100days}$ | 0.03 (-0.02, 0.07) | -0.05 (-0.06, -0.03) | 0 (-0.02, 0.03) |
| | $\Delta_{0\rightarrow 100days, Cohort}$ – $\Delta_{0\rightarrow 100days, Exposed}$ | Ref | **-0.07 (-0.12, -0.02)** | -0.03 (-0.08, 0.03) |
| 18 to 39 years | $Time_{0,Cohort}$ | 29.6 (28.9, 30.2) | 29.2 (28.8, 29.7) | 29.3 (28.6, 29.9) |
| | $Time_{0,Cohort}$ - $Time_{0,Exposed}$ | Ref | **-0.32 (-0.42, -0.22)** | **-0.30 (-0.42, -0.17)** |
| | $\Delta_{0\rightarrow 100days}$ | 0.06 (0.01, 0.11) | 0.07 (0.05, 0.1) | 0.09 (0.06, 0.12) |
| | $\Delta_{0\rightarrow 100days, Cohort}$ – $\Delta_{0\rightarrow 100days, Exposed}$ | Ref | 0.01 (-0.04, 0.07) | 0.04 (-0.02, 0.09) |
| 40 to 64 years | $Time_{0,Cohort}$ | 29.8 (29.1, 30.4) | 29.7 (29.4, 30.1) | 29.5 (28.8, 30.1) |
| | $Time_{0,Cohort}$ - $Time_{0,Exposed}$ | Ref | -0.04 (-0.12, 0.04) | **-0.30 (-0.41, -0.20)** |
| | $\Delta_{0\rightarrow 100days}$ | -0.01 (-0.05, 0.04) | -0.04 (-0.06, -0.03) | 0.02 (0, 0.04) |
| | $\Delta_{0\rightarrow 100days, Cohort}$ – $\Delta_{0\rightarrow 100days, Exposed}$ | Ref | -0.04 (-0.08, 0.01) | 0.03 (-0.02, 0.08) |
| 65 and older | $Time_{0,Cohort}$ | 29.6 (29.1, 30.1) | 29.7 (29.4, 29.9) | 29.5 (29, 30) |
| | $Time_{0,Cohort}$ - $Time_{0,Exposed}$ | Ref | 0.09 (-0.01, 0.18) | -0.09 (-0.22, 0.03) |
| | $\Delta_{0\rightarrow 100days}$ | -0.05 (-0.1, 0.01) | -0.09 (-0.11, -0.08) | -0.01 (-0.04, 0.01) |
| | $\Delta_{0\rightarrow 100days, Cohort}$ – $\Delta_{0\rightarrow 100days, Exposed}$ | Ref | -0.05 (-0.1, 0.01) | 0.03 (-0.02, 0.09) |
| Hispanic | $Time_{0,Cohort}$ | 29.4 (29.1, 29.7) | 29.3 (28.6, 30) | 29.4 (29, 29.7) |
| | $Time_{0,Cohort}$ - $Time_{0,Exposed}$ | Ref | -0.06 (-0.17, 0.05) | -0.01 (-0.16, 0.13) |
| | $\Delta_{0\rightarrow 100days}$ | 0.03 (-0.02, 0.08) | -0.02 (-0.05, 0.01) | -0.02 (-0.05, 0.02) |
| | $\Delta_{0\rightarrow 100days, Cohort}$ – $\Delta_{0\rightarrow 100days, Exposed}$ | Ref | -0.05 (-0.11, 0.01) | -0.05 (-0.11, 0.01) |
| Non-Hispanic White | $Time_{0,Cohort}$ | 29.4 (28.7, 30) | 29.3 (28.9, 29.6) | 29 (28.3, 29.8) |
| | $Time_{0,Cohort}$ - $Time_{0,Exposed}$ | Ref | **-0.08 (-0.16, -0.01)** | **-0.32 (-0.42, -0.23)** |
| | $\Delta_{0\rightarrow 100days}$ | -0.03 (-0.07, 0.01) | -0.04 (-0.06, -0.03) | 0.04 (0.02, 0.06) |

| | | | | |
|---|---|---|---|---|
| | $\Delta_{0\rightarrow 100\text{days, Cohort}}$ – $\Delta_{0\rightarrow 100\text{days, Exposed}}$ | Ref | -0.01 (-0.05, 0.03) | **0.07 (0.03, 0.11)** |
| Non-Hispanic Black | $\text{Time}_{0,\text{Cohort}}$ | 29.5 (28.9, 30.2) | 29.4 (28.7, 30) | 29.2 (28.5, 29.9) |
| | $\text{Time}_{0,\text{Cohort}}$ - $\text{Time}_{0,\text{Exposed}}$ | Ref | **-0.13 (-0.25, -0.02)** | **-0.34 (-0.48, -0.19)** |
| | $\Delta_{0\rightarrow 100\text{days}}$ | 0.01 (-0.05, 0.07) | -0.03 (-0.06, -0.01) | 0.05 (0.01, 0.08) |
| | $\Delta_{0\rightarrow 100\text{days, Cohort}}$ – $\Delta_{0\rightarrow 100\text{days, Exposed}}$ | Ref | -0.04 (-0.11, 0.02) | 0.04 (-0.03, 0.1) |
| Not Hospitalized | $\text{Time}_{0,\text{Cohort}}$ | 29.7 (29.4, 30.1) | 29.7 (29.5, 29.9) | 29.5 (29.2, 29.8) |
| | $\text{Time}_{0,\text{Cohort}}$ - $\text{Time}_{0,\text{Exposed}}$ | Ref | -0.04 (-0.09, 0.01) | **-0.26 (-0.33, -0.2)** |
| | $\Delta_{0\rightarrow 100\text{days}}$ | 0 (-0.03, 0.02) | -0.04 (-0.05, -0.03) | 0.03 (0.01, 0.04) |
| | $\Delta_{0\rightarrow 100\text{days, Cohort}}$ – $\Delta_{0\rightarrow 100\text{days, Exposed}}$ | Ref | -0.04 (-0.07, -0.01) | **0.03 (0, 0.06)** |
| Hospitalized | $\text{Time}_{0,\text{Cohort}}$ | 29.2 (28.4, 29.9) | 29 (28.7, 29.3) | 29.1 (27.8, 30.3) |
| | $\text{Time}_{0,\text{Cohort}}$ - $\text{Time}_{0,\text{Exposed}}$ | Ref | **-0.14 (-0.26, -0.03)** | **-0.09 (-0.31, 0.13)** |
| | $\Delta_{0\rightarrow 100\text{days}}$ | 0.01 (-0.05, 0.08) | -0.02 (-0.04, 0.01) | 0.05 (-0.02, 0.11) |
| | $\Delta_{0\rightarrow 100\text{days, Cohort}}$ – $\Delta_{0\rightarrow 100\text{days, Exposed}}$ | Ref | -0.03 (-0.1, 0.04) | 0.04 (-0.06, 0.13) |

**Supplementary Table 6. Modeled marginal estimates of changes in body mass index during follow-up by exposure cohort among those with 100 days of follow-up and at least two BMI encounters, n = 1,103,882 observations from 124,934 patients**

| | | **Exposed** | **Unexposed** | **Historical** |
|---|---|---|---|---|
| Overall | $Time_{0,Cohort}$ | 29.4 (29, 29.9) | 29.4 (29.2, 29.6) | 29.1 (28.7, 29.5) |
| | $Time_{0,Cohort}$ - $Time_{0,Exposed}$ | Ref | -0.06 (-0.13, 0.01) | **-0.29 (-0.37, -0.21)** |
| | $\Delta_{0\rightarrow 100days}$ | 0 (-0.03, 0.03) | -0.04 (-0.05, -0.03) | 0.03 (0.01, 0.04) |
| | $\Delta_{0\rightarrow 100days, Cohort}$ – $\Delta_{0\rightarrow 100days, Exposed}$ | Ref | -0.03 (-0.07, 0) | 0.03 (0, 0.06) |
| Female | $Time_{0,Cohort}$ | 29.9 (29.3, 30.4) | 29.7 (29.4, 30) | 29.5 (29, 29.9) |
| | $Time_{0,Cohort}$ - $Time_{0,Exposed}$ | Ref | **-0.16 (-0.25, -0.08)** | **-0.41 (-0.51, -0.31)** |
| | $\Delta_{0\rightarrow 100days}$ | -0.02 (-0.06, 0.01) | -0.03 (-0.05, -0.02) | 0.05 (0.03, 0.07) |
| | $\Delta_{0\rightarrow 100days, Cohort}$ – $\Delta_{0\rightarrow 100days, Exposed}$ | Ref | -0.01 (-0.05, 0.03) | 0.07 (0.03, 0.11) |
| Male | $Time_{0,Cohort}$ | 29.6 (28.9, 30.3) | 29.7 (29.4, 30) | 29.6 (29, 30.1) |
| | $Time_{0,Cohort}$ - $Time_{0,Exposed}$ | Ref | 0.07 (-0.04, 0.18) | -0.08 (-0.21, 0.05) |
| | $\Delta_{0\rightarrow 100days}$ | 0.03 (-0.02, 0.07) | -0.05 (-0.07, -0.03) | 0 (-0.02, 0.03) |
| | $\Delta_{0\rightarrow 100days, Cohort}$ – $\Delta_{0\rightarrow 100days, Exposed}$ | Ref | **-0.07 (-0.12, -0.02)** | **-0.03 (-0.08, 0.03)** |
| 18 to 39 years | $Time_{0,Cohort}$ | 29.6 (28.8, 30.5) | 29.3 (28.8, 29.9) | 29.3 (28.5, 30.1) |
| | $Time_{0,Cohort}$ - $Time_{0,Exposed}$ | Ref | **-0.33 (-0.46, -0.21)** | **-0.33 (-0.48, -0.18)** |
| | $\Delta_{0\rightarrow 100days}$ | 0.06 (0.01, 0.11) | 0.07 (0.05, 0.1) | 0.09 (0.06, 0.12) |
| | $\Delta_{0\rightarrow 100days, Cohort}$ – $\Delta_{0\rightarrow 100days, Exposed}$ | Ref | 0.01 (-0.04, 0.07) | 0.04 (-0.02, 0.09) |
| 40 to 64 years | $Time_{0,Cohort}$ | 29.8 (29, 30.7) | 29.8 (29.4, 30.2) | 29.5 (28.8, 30.2) |
| | $Time_{0,Cohort}$ - $Time_{0,Exposed}$ | Ref | -0.03 (-0.13, 0.07) | **-0.31 (-0.43, -0.19)** |
| | $\Delta_{0\rightarrow 100days}$ | -0.01 (-0.05, 0.04) | -0.05 (-0.06, -0.03) | 0.02 (0, 0.04) |
| | $\Delta_{0\rightarrow 100days, Cohort}$ – $\Delta_{0\rightarrow 100days, Exposed}$ | Ref | -0.04 (-0.08, 0.01) | 0.03 (-0.02, 0.08) |
| 65 and older | $Time_{0,Cohort}$ | 29.8 (29.1, 30.4) | 29.8 (29.5, 30.1) | 29.6 (29.1, 30.1) |
| | $Time_{0,Cohort}$ - $Time_{0,Exposed}$ | Ref | 0.07 (-0.06, 0.19) | **-0.15 (-0.3, 0.00)** |
| | $\Delta_{0\rightarrow 100days}$ | -0.05 (-0.1, 0) | -0.09 (-0.11, -0.08) | -0.01 (-0.04, 0.01) |
| | $\Delta_{0\rightarrow 100days, Cohort}$ – $\Delta_{0\rightarrow 100days, Exposed}$ | Ref | -0.05 (-0.1, 0.01) | 0.03 (-0.02, 0.09) |
| Hispanic | $Time_{0,Cohort}$ | 29.5 (29.1, 29.9) | 29.4 (28.6, 30.3) | 29.4 (29, 29.9) |
| | $Time_{0,Cohort}$ - $Time_{0,Exposed}$ | Ref | -0.01 (-0.15, 0.13) | -0.02 (-0.19, 0.15) |

| | | | | |
|---|---|---|---|---|
| | $\Delta_{0 \rightarrow 100days}$ | 0.03 (-0.03, 0.08) | -0.02 (-0.05, 0.01) | -0.02 (-0.05, 0.02) |
| | $\Delta_{0 \rightarrow 100days, Cohort}$ – $\Delta_{0 \rightarrow 100days, Exposed}$ | Ref | -0.05 (-0.11, 0.01) | -0.04 (-0.11, 0.02) |
| Non-Hispanic White | $Time_{0,Cohort}$ | 29.5 (28.7, 30.2) | 29.4 (29, 29.8) | 29.1 (28.3, 30) |
| | $Time_{0,Cohort}$ - $Time_{0,Exposed}$ | Ref | **-0.09 (-0.19, 0)** | **-0.33 (-0.44, -0.22)** |
| | $\Delta_{0 \rightarrow 100days}$ | -0.03 (-0.07, 0.01) | -0.05 (-0.06, -0.03) | 0.04 (0.02, 0.06) |
| | $\Delta_{0 \rightarrow 100days, Cohort}$ – $\Delta_{0 \rightarrow 100days, Exposed}$ | Ref | -0.01 (-0.06, 0.03) | **0.07 (0.03, 0.12)** |
| Non-Hispanic Black | $Time_{0,Cohort}$ | 29.6 (28.9, 30.3) | 29.5 (28.6, 30.3) | 29.2 (28.5, 30) |
| | $Time_{0,Cohort}$ - $Time_{0,Exposed}$ | Ref | **-0.13 (-0.27, 0)** | **-0.39 (-0.55, -0.22)** |
| | $\Delta_{0 \rightarrow 100days}$ | 0.01 (-0.05, 0.07) | -0.03 (-0.06, -0.01) | 0.05 (0.01, 0.08) |
| | $\Delta_{0 \rightarrow 100days, Cohort}$ – $\Delta_{0 \rightarrow 100days, Exposed}$ | Ref | -0.04 (-0.1, 0.02) | 0.04 (-0.03, 0.1) |
| Not Hospitalized | $Time_{0,Cohort}$ | 29.8 (29.4, 30.2) | 29.8 (29.5, 30) | 29.5 (29.2, 29.9) |
| | $Time_{0,Cohort}$ - $Time_{0,Exposed}$ | Ref | **-0.06 (-0.13, 0)** | **-0.3 (-0.37, -0.22)** |
| | $\Delta_{0 \rightarrow 100days}$ | -0.01 (-0.03, 0.02) | -0.04 (-0.06, -0.03) | 0.03 (0.01, 0.04) |
| | $\Delta_{0 \rightarrow 100days, Cohort}$ – $\Delta_{0 \rightarrow 100days, Exposed}$ | Ref | **-0.04 (-0.07, -0.01)** | **0.03 (0, 0.06)** |
| Hospitalized | $Time_{0,Cohort}$ | 29.3 (28.3, 30.2) | 29.2 (28.8, 29.6) | 29.1 (27.7, 30.5) |
| | $Time_{0,Cohort}$ - $Time_{0,Exposed}$ | Ref | -0.08 (-0.23, 0.07) | -0.15 (-0.41, 0.10) |
| | $\Delta_{0 \rightarrow 100days}$ | 0.01 (-0.06, 0.07) | -0.02 (-0.04, 0) | 0.05 (-0.02, 0.12) |
| | $\Delta_{0 \rightarrow 100days, Cohort}$ – $\Delta_{0 \rightarrow 100days, Exposed}$ | Ref | -0.03 (-0.1, 0.04) | 0.04 (-0.05, 0.14) |

**Supplementary Table 7. Modeled marginal estimates of changes in body mass index during follow-up by exposure cohort after adjusting for number and type of encounters between BMI measurements, n = 1,230,631 observations from 194,792 patients**

| | | **Exposed** | **Unexposed** | **Historical** |
|---|---|---|---|---|
| Overall | $Time_{0,Cohort}$ | 29.3 (29, 29.7) | 29.3 (29.1, 29.4) | 29.1 (28.7, 29.4) |
| | $Time_{0,Cohort}$ - $Time_{0,Exposed}$ | Ref | **-0.07 (-0.12, -0.01)** | **-0.27 (-0.34, -0.2)** |
| | $\Delta_{0\rightarrow 100days}$ | 0 (-0.03, 0.03) | -0.04 (-0.05, -0.02) | 0.03 (0.02, 0.04) |
| | $\Delta_{0\rightarrow 100days, Cohort}$ – $\Delta_{0\rightarrow 100days, Exposed}$ | Ref | **-0.04 (-0.06, -0.01)** | **0.03 (0, 0.06)** |
| Female | $Time_{0,Cohort}$ | 29.8 (29.3, 30.2) | 29.6 (29.4, 29.8) | 29.4 (29, 29.8) |
| | $Time_{0,Cohort}$ - $Time_{0,Exposed}$ | Ref | **-0.16 (-0.23, -0.09)** | **-0.36 (-0.45, -0.27)** |
| | $\Delta_{0\rightarrow 100days}$ | -0.02 (-0.05, 0.01) | -0.03 (-0.04, -0.01) | 0.05 (0.03, 0.07) |
| | $\Delta_{0\rightarrow 100days, Cohort}$ – $\Delta_{0\rightarrow 100days, Exposed}$ | Ref | -0.01 (-0.05, 0.03) | **0.07 (0.03, 0.11)** |
| Male | $Time_{0,Cohort}$ | 29.5 (29, 30.1) | 29.6 (29.4, 29.9) | 29.5 (28.9, 30) |
| | $Time_{0,Cohort}$ - $Time_{0,Exposed}$ | Ref | 0.07 (-0.02, 0.16) | -0.09 (-0.2, 0.03) |
| | $\Delta_{0\rightarrow 100days}$ | 0.03 (-0.02, 0.07) | -0.05 (-0.06, -0.03) | 0 (-0.02, 0.03) |
| | $\Delta_{0\rightarrow 100days, Cohort}$ – $\Delta_{0\rightarrow 100days, Exposed}$ | Ref | -0.07 (-0.12, -0.02) | -0.03 (-0.08, 0.03) |
| 18 to 39 years | $Time_{0,Cohort}$ | 29.6 (28.9, 30.2) | 29.2 (28.8, 29.7) | 29.3 (28.6, 29.9) |
| | $Time_{0,Cohort}$ - $Time_{0,Exposed}$ | Ref | -0.32 (-0.42, -0.22) | -0.3 (-0.43, -0.17) |
| | $\Delta_{0\rightarrow 100days}$ | 0.06 (0.01, 0.11) | 0.07 (0.05, 0.1) | 0.09 (0.06, 0.12) |
| | $\Delta_{0\rightarrow 100days, Cohort}$ – $\Delta_{0\rightarrow 100days, Exposed}$ | Ref | 0.02 (-0.04, 0.07) | 0.03 (-0.02, 0.09) |
| 40 to 64 years | $Time_{0,Cohort}$ | 29.8 (29.1, 30.4) | 29.7 (29.4, 30.1) | 29.5 (28.8, 30.1) |
| | $Time_{0,Cohort}$ - $Time_{0,Exposed}$ | Ref | -0.04 (-0.12, 0.04) | -0.3 (-0.41, -0.2) |
| | $\Delta_{0\rightarrow 100days}$ | -0.01 (-0.05, 0.04) | -0.04 (-0.06, -0.03) | 0.02 (0, 0.04) |
| | $\Delta_{0\rightarrow 100days, Cohort}$ – $\Delta_{0\rightarrow 100days, Exposed}$ | Ref | -0.04 (-0.08, 0.01) | 0.03 (-0.02, 0.07) |
| 65 and older | $Time_{0,Cohort}$ | 29.6 (29.1, 30.1) | 29.7 (29.4, 29.9) | 29.5 (29, 30) |
| | $Time_{0,Cohort}$ - $Time_{0,Exposed}$ | Ref | 0.09 (-0.01, 0.19) | -0.1 (-0.22, 0.03) |
| | $\Delta_{0\rightarrow 100days}$ | -0.04 (-0.09, 0.01) | -0.09 (-0.11, -0.08) | -0.01 (-0.04, 0.01) |
| | $\Delta_{0\rightarrow 100days, Cohort}$ – $\Delta_{0\rightarrow 100days, Exposed}$ | Ref | -0.05 (-0.1, 0.01) | 0.03 (-0.02, 0.09) |
| Hispanic | $Time_{0,Cohort}$ | 29.4 (29.1, 29.7) | 29.3 (28.6, 30) | 29.4 (29, 29.7) |
| | $Time_{0,Cohort}$ - $Time_{0,Exposed}$ | Ref | -0.06 (-0.17, 0.05) | -0.01 (-0.16, 0.13) |

| | | | | |
|---|---|---|---|---|
| | $\Delta_{0\rightarrow 100days}$ | 0.03 (-0.02, 0.08) | -0.02 (-0.05, 0.01) | -0.02 (-0.05, 0.02) |
| | $\Delta_{0\rightarrow 100days, Cohort}$ – $\Delta_{0\rightarrow 100days, Exposed}$ | Ref | -0.05 (-0.11, 0.01) | -0.05 (-0.11, 0.01) |
| Non-Hispanic White | $Time_{0,Cohort}$ | 29.4 (28.7, 30) | 29.3 (28.9, 29.6) | 29 (28.3, 29.8) |
| | $Time_{0,Cohort}$ - $Time_{0,Exposed}$ | Ref | -0.08 (-0.15, 0) | -0.32 (-0.42, -0.23) |
| | $\Delta_{0\rightarrow 100days}$ | -0.03 (-0.07, 0.01) | -0.04 (-0.06, -0.03) | 0.04 (0.02, 0.06) |
| | $\Delta_{0\rightarrow 100days, Cohort}$ – $\Delta_{0\rightarrow 100days, Exposed}$ | Ref | -0.01 (-0.05, 0.03) | 0.07 (0.03, 0.11) |
| Non-Hispanic Black | $Time_{0,Cohort}$ | 29.5 (28.9, 30.2) | 29.4 (28.7, 30) | 29.2 (28.5, 29.9) |
| | $Time_{0,Cohort}$ - $Time_{0,Exposed}$ | Ref | -0.13 (-0.24, -0.02) | -0.34 (-0.48, -0.19) |
| | $\Delta_{0\rightarrow 100days}$ | 0.01 (-0.05, 0.07) | -0.03 (-0.06, -0.01) | 0.05 (0.02, 0.08) |
| | $\Delta_{0\rightarrow 100days, Cohort}$ – $\Delta_{0\rightarrow 100days, Exposed}$ | Ref | -0.04 (-0.11, 0.02) | 0.04 (-0.03, 0.1) |
| Not Hospitalized | $Time_{0,Cohort}$ | 29.7 (29.4, 30.1) | 29.7 (29.5, 29.9) | 29.5 (29.2, 29.8) |
| | $Time_{0,Cohort}$ - $Time_{0,Exposed}$ | Ref | -0.04 (-0.09, 0.01) | -0.27 (-0.33, -0.2) |
| | $\Delta_{0\rightarrow 100days}$ | 0 (-0.03, 0.02) | -0.04 (-0.05, -0.03) | 0.03 (0.01, 0.04) |
| | $\Delta_{0\rightarrow 100days, Cohort}$ – $\Delta_{0\rightarrow 100days, Exposed}$ | Ref | -0.04 (-0.07, -0.01) | 0.03 (0, 0.06) |
| Hospitalized | $Time_{0,Cohort}$ | 29.2 (28.4, 29.9) | 29 (28.7, 29.3) | 29.1 (27.8, 30.4) |
| | $Time_{0,Cohort}$ - $Time_{0,Exposed}$ | Ref | -0.14 (-0.25, -0.02) | -0.09 (-0.31, 0.13) |
| | $\Delta_{0\rightarrow 100days}$ | 0.01 (-0.05, 0.08) | -0.02 (-0.04, 0.01) | 0.05 (-0.02, 0.12) |
| | $\Delta_{0\rightarrow 100days, Cohort}$ – $\Delta_{0\rightarrow 100days, Exposed}$ | Ref | -0.03 (-0.1, 0.04) | 0.04 (-0.06, 0.13) |

**Supplementary Table 8. Modeled marginal estimates of changes in body mass index during follow-up by exposure cohort after adjusting for COVID-19 cases and test positivity rate, n = 1,230,631 observations from 194,792 patients**

| | | **Exposed** | **Unexposed** | **Historical** |
|---|---|---|---|---|
| Overall | $Time_{0,Cohort}$ | 29.3 (29, 29.7) | 29.3 (29.1, 29.4) | 29.1 (28.7, 29.4) |
| | $Time_{0,Cohort}$ - $Time_{0,Exposed}$ | Ref | **-0.07 (-0.12, -0.01)** | **-0.27 (-0.34, -0.2)** |
| | $\Delta_{0\rightarrow 100days}$ | 0 (-0.03, 0.03) | -0.03 (-0.05, -0.02) | 0.03 (0.01, 0.04) |
| | $\Delta_{0\rightarrow 100days,\ Cohort}$ – $\Delta_{0\rightarrow 100days,\ Exposed}$ | Ref | -0.04 (-0.07, -0.01) | 0.03 (0, 0.06) |
| Female | $Time_{0,Cohort}$ | 29.8 (29.3, 30.2) | 29.6 (29.4, 29.8) | 29.4 (29, 29.8) |
| | $Time_{0,Cohort}$ - $Time_{0,Exposed}$ | Ref | -0.16 (-0.23, -0.1) | -0.36 (-0.45, -0.28) |
| | $\Delta_{0\rightarrow 100days}$ | -0.02 (-0.05, 0.02) | -0.03 (-0.04, -0.01) | 0.05 (0.03, 0.07) |
| | $\Delta_{0\rightarrow 100days,\ Cohort}$ – $\Delta_{0\rightarrow 100days,\ Exposed}$ | Ref | -0.01 (-0.05, 0.03) | 0.07 (0.03, 0.11) |
| Male | $Time_{0,Cohort}$ | 29.5 (29, 30.1) | 29.6 (29.4, 29.9) | 29.5 (28.9, 30) |
| | $Time_{0,Cohort}$ - $Time_{0,Exposed}$ | Ref | 0.07 (-0.02, 0.16) | -0.09 (-0.2, 0.02) |
| | $\Delta_{0\rightarrow 100days}$ | 0.03 (-0.02, 0.08) | -0.04 (-0.06, -0.03) | 0 (-0.02, 0.03) |
| | $\Delta_{0\rightarrow 100days,\ Cohort}$ – $\Delta_{0\rightarrow 100days,\ Exposed}$ | Ref | -0.07 (-0.12, -0.02) | -0.03 (-0.08, 0.03) |
| 18 to 39 years | $Time_{0,Cohort}$ | 29.6 (28.9, 30.2) | 29.2 (28.8, 29.7) | 29.3 (28.6, 29.9) |
| | $Time_{0,Cohort}$ - $Time_{0,Exposed}$ | Ref | -0.32 (-0.42, -0.23) | -0.3 (-0.43, -0.18) |
| | $\Delta_{0\rightarrow 100days}$ | 0.06 (0.01, 0.11) | 0.07 (0.05, 0.1) | 0.09 (0.06, 0.12) |
| | $\Delta_{0\rightarrow 100days,\ Cohort}$ – $\Delta_{0\rightarrow 100days,\ Exposed}$ | Ref | 0.01 (-0.04, 0.07) | 0.03 (-0.02, 0.09) |
| 40 to 64 years | $Time_{0,Cohort}$ | 29.8 (29.1, 30.4) | 29.7 (29.4, 30.1) | 29.5 (28.8, 30.1) |
| | $Time_{0,Cohort}$ - $Time_{0,Exposed}$ | Ref | -0.04 (-0.12, 0.04) | -0.31 (-0.41, -0.2) |
| | $\Delta_{0\rightarrow 100days}$ | -0.01 (-0.05, 0.04) | -0.04 (-0.06, -0.02) | 0.02 (0, 0.04) |
| | $\Delta_{0\rightarrow 100days,\ Cohort}$ – $\Delta_{0\rightarrow 100days,\ Exposed}$ | Ref | -0.04 (-0.08, 0.01) | 0.03 (-0.02, 0.07) |
| 65 and older | $Time_{0,Cohort}$ | 29.6 (29.1, 30.1) | 29.7 (29.4, 29.9) | 29.5 (29, 30) |
| | $Time_{0,Cohort}$ - $Time_{0,Exposed}$ | Ref | 0.09 (-0.01, 0.18) | -0.1 (-0.22, 0.03) |
| | $\Delta_{0\rightarrow 100days}$ | -0.04 (-0.09, 0.01) | -0.09 (-0.11, -0.07) | -0.01 (-0.04, 0.01) |
| | $\Delta_{0\rightarrow 100days,\ Cohort}$ – $\Delta_{0\rightarrow 100days,\ Exposed}$ | Ref | -0.05 (-0.1, 0.01) | 0.03 (-0.03, 0.09) |
| Hispanic | $Time_{0,Cohort}$ | 29.4 (29.1, 29.7) | 29.3 (28.6, 30) | 29.4 (29, 29.7) |
| | $Time_{0,Cohort}$ - $Time_{0,Exposed}$ | Ref | -0.06 (-0.17, 0.05) | -0.02 (-0.17, 0.13) |

| | | | | |
|---|---|---|---|---|
| | $\Delta_{0\rightarrow 100days}$ | 0.03 (-0.02, 0.08) | -0.02 (-0.05, 0.01) | -0.02 (-0.05, 0.02) |
| | $\Delta_{0\rightarrow 100days, Cohort}$ – $\Delta_{0\rightarrow 100days, Exposed}$ | Ref | -0.05 (-0.11, 0.01) | -0.05 (-0.11, 0.01) |
| Non-Hispanic White | $Time_{0,Cohort}$ | 29.4 (28.7, 30) | 29.3 (28.9, 29.6) | 29 (28.3, 29.8) |
| | $Time_{0,Cohort}$ - $Time_{0,Exposed}$ | Ref | -0.08 (-0.16, -0.01) | -0.33 (-0.42, -0.23) |
| | $\Delta_{0\rightarrow 100days}$ | -0.03 (-0.07, 0.01) | -0.04 (-0.06, -0.03) | 0.04 (0.02, 0.06) |
| | $\Delta_{0\rightarrow 100days, Cohort}$ – $\Delta_{0\rightarrow 100days, Exposed}$ | Ref | -0.01 (-0.05, 0.03) | 0.07 (0.02, 0.11) |
| Non-Hispanic Black | $Time_{0,Cohort}$ | 29.5 (28.9, 30.2) | 29.4 (28.7, 30) | 29.2 (28.4, 29.9) |
| | $Time_{0,Cohort}$ - $Time_{0,Exposed}$ | Ref | -0.13 (-0.25, -0.02) | -0.34 (-0.49, -0.19) |
| | $\Delta_{0\rightarrow 100days}$ | 0.01 (-0.05, 0.07) | -0.03 (-0.06, -0.01) | 0.05 (0.01, 0.08) |
| | $\Delta_{0\rightarrow 100days, Cohort}$ – $\Delta_{0\rightarrow 100days, Exposed}$ | Ref | -0.04 (-0.11, 0.02) | 0.03 (-0.03, 0.1) |
| Not Hospitalized | $Time_{0,Cohort}$ | 29.7 (29.4, 30.1) | 29.7 (29.5, 29.9) | 29.5 (29.1, 29.8) |
| | $Time_{0,Cohort}$ - $Time_{0,Exposed}$ | Ref | -0.04 (-0.1, 0.01) | -0.27 (-0.34, -0.21) |
| | $\Delta_{0\rightarrow 100days}$ | 0 (-0.02, 0.02) | -0.04 (-0.05, -0.03) | 0.03 (0.01, 0.04) |
| | $\Delta_{0\rightarrow 100days, Cohort}$ – $\Delta_{0\rightarrow 100days, Exposed}$ | Ref | -0.04 (-0.07, -0.01) | 0.03 (0, 0.06) |
| Hospitalized | $Time_{0,Cohort}$ | 29.2 (28.4, 29.9) | 29 (28.7, 29.3) | 29.1 (27.8, 30.3) |
| | $Time_{0,Cohort}$ - $Time_{0,Exposed}$ | Ref | -0.14 (-0.26, -0.03) | -0.1 (-0.32, 0.12) |
| | $\Delta_{0\rightarrow 100days}$ | 0.02 (-0.05, 0.08) | -0.01 (-0.04, 0.01) | 0.05 (-0.02, 0.11) |
| | $\Delta_{0\rightarrow 100days, Cohort}$ – $\Delta_{0\rightarrow 100days, Exposed}$ | Ref | -0.03 (-0.1, 0.04) | 0.03 (-0.06, 0.13) |

**Supplementary Table 9. Modeled marginal estimates of changes in body mass index during follow-up by exposure cohort after adjusting for laboratory parameters with multiple imputation, n = 1,230,631 observations from 194,792 patients**

| | | **Exposed** | **Unexposed** | **Historical** |
|---|---|---|---|---|
| Overall | $Time_{0,Cohort}$ | 29.4 | 29.3 | 29.1 |
| | $Time_{0,Cohort}$ - $Time_{0,Exposed}$ | 0 | -0.06 (-0.11, 0) | -0.26 (-0.33, -0.2) |
| | $\Delta_{0\rightarrow 100days}$ | 0 (-0.03, 0.03) | -0.04 (-0.05, -0.02) | 0.03 (0.01, 0.04) |
| | $\Delta_{0\rightarrow 100days,\ Cohort}$ – $\Delta_{0\rightarrow 100days,\ Exposed}$ | 0 | -0.04 (-0.06, -0.01) | 0.03 (0, 0.06) |
| Female | $Time_{0,Cohort}$ | 29.8 | 29.6 | 29.4 |
| | $Time_{0,Cohort}$ - $Time_{0,Exposed}$ | 0 | -0.15 (-0.22, -0.09) | -0.36 (-0.45, -0.27) |
| | $\Delta_{0\rightarrow 100days}$ | -0.02 (-0.05, 0.01) | -0.03 (-0.05, -0.02) | 0.05 (0.03, 0.07) |
| | $\Delta_{0\rightarrow 100days,\ Cohort}$ – $\Delta_{0\rightarrow 100days,\ Exposed}$ | 0 | -0.01 (-0.05, 0.03) | 0.07 (0.03, 0.11) |
| Male | $Time_{0,Cohort}$ | 29.6 | 29.6 | 29.5 |
| | $Time_{0,Cohort}$ - $Time_{0,Exposed}$ | 0 | 0.08 (-0.01, 0.17) | -0.09 (-0.2, 0.02) |
| | $\Delta_{0\rightarrow 100days}$ | 0.03 (-0.02, 0.07) | -0.05 (-0.06, -0.03) | 0 (-0.02, 0.03) |
| | $\Delta_{0\rightarrow 100days,\ Cohort}$ – $\Delta_{0\rightarrow 100days,\ Exposed}$ | 0 | -0.07 (-0.12, -0.02) | -0.03 (-0.08, 0.03) |
| 18 to 39 years | $Time_{0,Cohort}$ | 29.6 | 29.3 | 29.3 |
| | $Time_{0,Cohort}$ - $Time_{0,Exposed}$ | 0 | -0.31 (-0.41, -0.21) | -0.3 (-0.42, -0.17) |
| | $\Delta_{0\rightarrow 100days}$ | 0.06 (0.01, 0.11) | 0.07 (0.05, 0.1) | 0.09 (0.06, 0.12) |
| | $\Delta_{0\rightarrow 100days,\ Cohort}$ – $\Delta_{0\rightarrow 100days,\ Exposed}$ | 0 | 0.01 (-0.04, 0.07) | 0.04 (-0.02, 0.09) |
| 40 to 64 years | $Time_{0,Cohort}$ | 29.8 | 29.8 | 29.5 |
| | $Time_{0,Cohort}$ - $Time_{0,Exposed}$ | 0 | -0.03 (-0.11, 0.05) | -0.3 (-0.41, -0.19) |
| | $\Delta_{0\rightarrow 100days}$ | -0.01 (-0.05, 0.04) | -0.04 (-0.06, -0.03) | 0.02 (0, 0.04) |
| | $\Delta_{0\rightarrow 100days,\ Cohort}$ – $\Delta_{0\rightarrow 100days,\ Exposed}$ | 0 | -0.04 (-0.08, 0.01) | 0.03 (-0.02, 0.08) |
| 65 and older | $Time_{0,Cohort}$ | 29.6 | 29.7 | 29.5 |
| | $Time_{0,Cohort}$ - $Time_{0,Exposed}$ | 0 | 0.1 (0, 0.2) | -0.09 (-0.21, 0.04) |
| | $\Delta_{0\rightarrow 100days}$ | -0.04 (-0.1, 0.01) | -0.09 (-0.11, -0.08) | -0.01 (-0.04, 0.01) |
| | $\Delta_{0\rightarrow 100days,\ Cohort}$ – $\Delta_{0\rightarrow 100days,\ Exposed}$ | 0 | -0.05 (-0.1, 0.01) | 0.03 (-0.02, 0.09) |
| Hispanic | $Time_{0,Cohort}$ | 29.4 | 29.3 | 29.4 |
| | $Time_{0,Cohort}$ - $Time_{0,Exposed}$ | 0 | -0.06 (-0.17, 0.05) | -0.02 (-0.16, 0.13) |

| | | | | |
|---|---|---|---|---|
| | $\Delta_{0\rightarrow 100days}$ | 0.03 (-0.02, 0.08) | -0.02 (-0.05, 0.01) | -0.02 (-0.05, 0.02) |
| | $\Delta_{0\rightarrow 100days, Cohort}$ – $\Delta_{0\rightarrow 100days, Exposed}$ | 0 | -0.05 (-0.11, 0.01) | -0.05 (-0.11, 0.01) |
| Non-Hispanic White | $Time_{0,Cohort}$ | 29.4 | 29.3 | 29.1 |
| | $Time_{0,Cohort}$ - $Time_{0,Exposed}$ | 0 | -0.07 (-0.15, 0) | -0.32 (-0.42, -0.23) |
| | $\Delta_{0\rightarrow 100days}$ | -0.03 (-0.07, 0.01) | -0.04 (-0.06, -0.03) | 0.04 (0.02, 0.06) |
| | $\Delta_{0\rightarrow 100days, Cohort}$ – $\Delta_{0\rightarrow 100days, Exposed}$ | 0 | -0.01 (-0.05, 0.03) | 0.07 (0.03, 0.11) |
| Non-Hispanic Black | $Time_{0,Cohort}$ | 29.5 | 29.4 | 29.2 |
| | $Time_{0,Cohort}$ - $Time_{0,Exposed}$ | 0 | -0.12 (-0.24, -0.01) | -0.33 (-0.48, -0.19) |
| | $\Delta_{0\rightarrow 100days}$ | 0.01 (-0.05, 0.07) | -0.03 (-0.06, -0.01) | 0.05 (0.01, 0.08) |
| | $\Delta_{0\rightarrow 100days, Cohort}$ – $\Delta_{0\rightarrow 100days, Exposed}$ | 0 | -0.04 (-0.11, 0.02) | 0.04 (-0.03, 0.1) |
| Not Hospitalized | $Time_{0,Cohort}$ | 29.8 | 29.8 | 29.5 |
| | $Time_{0,Cohort}$ - $Time_{0,Exposed}$ | 0 | -0.02 (-0.08, 0.03) | -0.26 (-0.32, -0.19) |
| | $\Delta_{0\rightarrow 100days}$ | 0 (-0.03, 0.02) | -0.04 (-0.05, -0.03) | 0.03 (0.01, 0.04) |
| | $\Delta_{0\rightarrow 100days, Cohort}$ – $\Delta_{0\rightarrow 100days, Exposed}$ | 0 | -0.04 (-0.07, -0.01) | 0.03 (0, 0.06) |
| Hospitalized | $Time_{0,Cohort}$ | 29.2 | 29.1 | 29.1 |
| | $Time_{0,Cohort}$ - $Time_{0,Exposed}$ | 0 | -0.13 (-0.24, -0.01) | -0.07 (-0.29, 0.15) |
| | $\Delta_{0\rightarrow 100days}$ | 0.01 (-0.05, 0.08) | -0.02 (-0.04, 0.01) | 0.05 (-0.02, 0.12) |
| | $\Delta_{0\rightarrow 100days, Cohort}$ – $\Delta_{0\rightarrow 100days, Exposed}$ | 0 | -0.03 (-0.1, 0.04) | 0.04 (-0.06, 0.13) |

Confidence intervals unavailable for predicted values at T=0 for all cohorts.

#### Supplementary Figure 1. Cohort selection

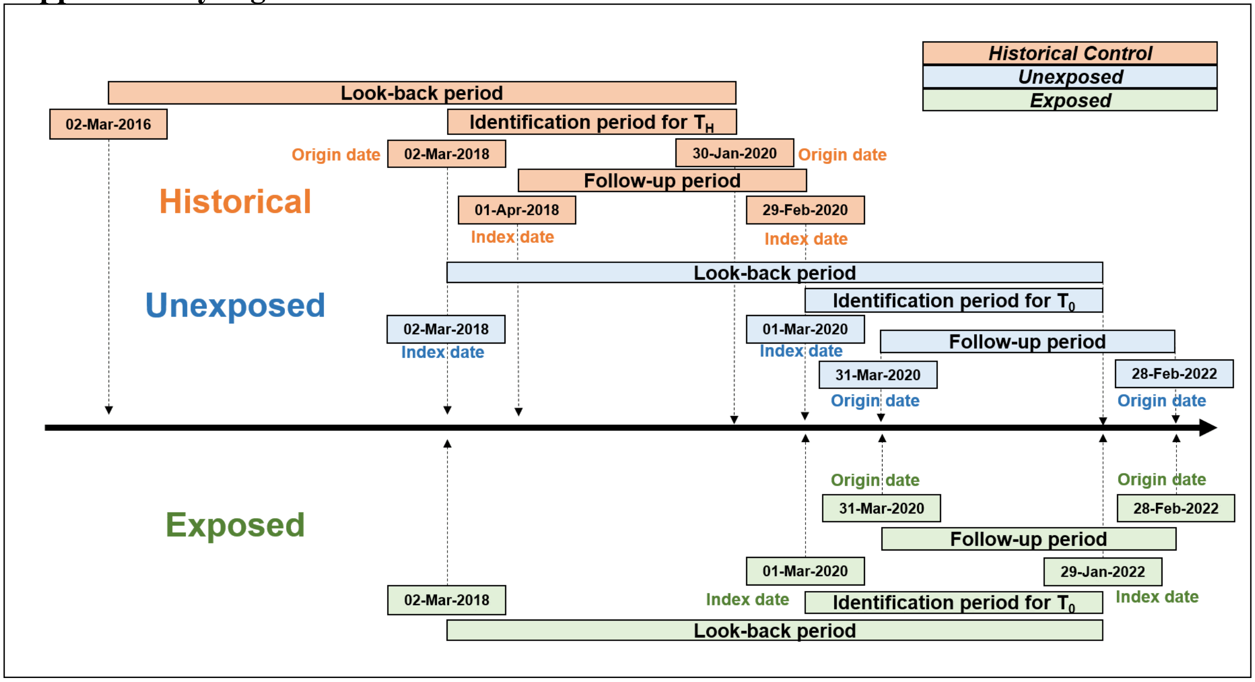

**Supplementary Figure 2. Flowchart of analytic sample selection**

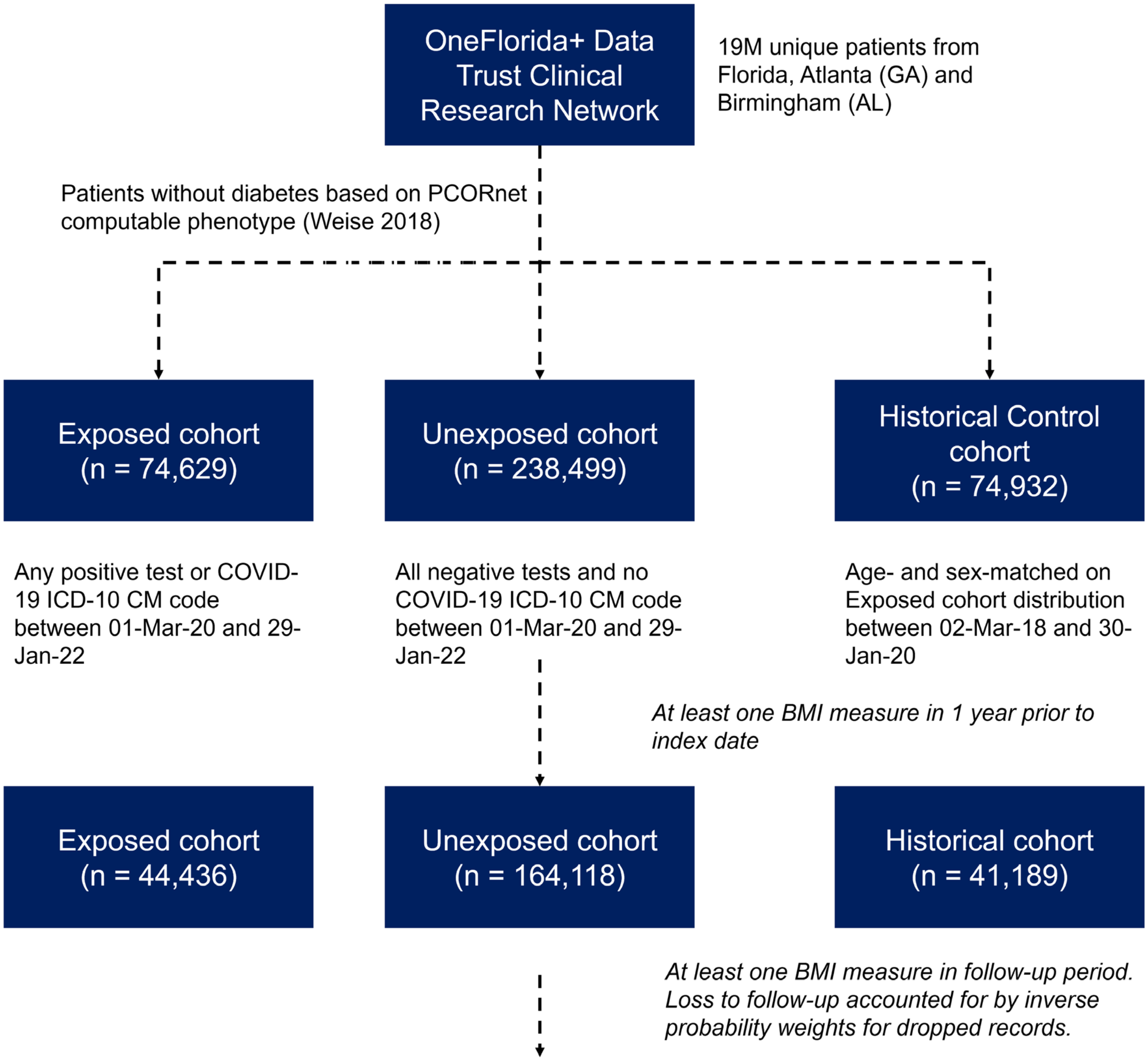

| | | Exposed | Unexposed | Historical |
|---|---|---|---|---|
| BMI | Unique patients | 32,384 | 127,142 | 35,266 |
| | Observations in follow-up | 188,060 | 809,483 | 233,088 |

**Supplementary Figure 3. Statistical analysis approach**

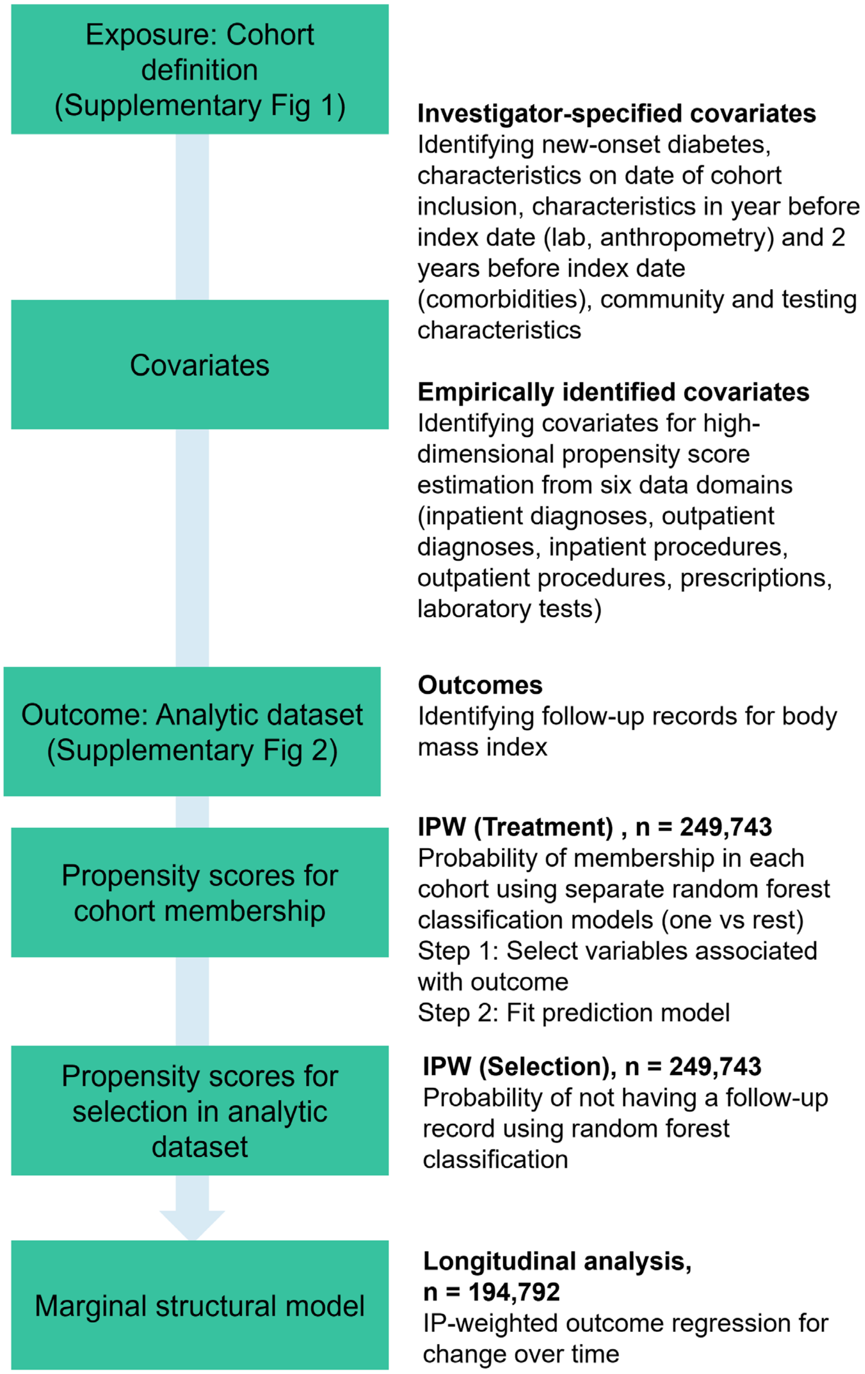

**Supplementary Methods. Detailed statistical methods**
***Step 1. Investigator specified covariates***

We extracted investigator specified covariates from the lookback period for each patient in the study sample (**Table SM1**). We imputed values as ‘no’ for those medications, comorbidities and encounter types if they were not present in the electronic health record for that patient. For other classes of nominal variables (health system, insurance category) we imputed missing values with the mode of the dataset. For continuous laboratory and anthropometry variables, missing data were single-imputed as the mean of available values across combinations of age, sex, race-ethnicity, health system, and exposure cohort..

**Table SM1. Investigator-specified covariates prior to outcome measurement**

| **Demographics** | **At or closest to index date** | **Within 1 year of index date** | **Within 2 years of index date** |
|---|---|---|---|
| Sex | Age and calendar month of admission | Smoking status (yes/no) [c] | Comorbidities using grouped ICD10-CM codes (yes/no: 6 variables: obesity, cardiovascular, cerebrovascular, hypertension, pulmonary, hyperlipidemia)[a] |
| Race/ethnicity (4 categories: NH White, NH Black, Hispanic, NH Other) | Health system (8 categories) [b] | Medication classes (yes/no: 5 variables; antidepressants, antipsychotics, antihypertensives, statins, immune suppressants) [a] | |
| | Primary and secondary insurance type (7 categories) [b] | Laboratory parameters (actual; HbA1c, Glucose, ALT, AST, SCreatinine, HDL, LDL) [c] | |
| | Hospitalization status (yes/no) and counts of encounters (hospitalization, not hospitalization) [a] | Anthropometry (actual; BMI, height, SBP) [c] | |
| | Hyperglycemia in blood glucose values between index date and origin date [c] | Times encounters were observed (counts; Hospitalization, Telehealth, Outpatient) [a] | |
| | | Times laboratory parameters were assessed (counts; Glucose, HDL, LDL, SCreatinine, ALT, AST, HbA1c, any lab visit) [a] | |

a Missing values set to ‘no’ ~ 0
b Missing values imputed using mode
c Missing values imputed using group mean (sex, race/ethnicity, age, health system)

***Step 2. Empirically identified covariates***

Recommendations for high dimensional propensity scores (Rassen 2023 *Pharmacoepidemiol Drug Saf.*) suggest categorizing available data dimensions to identify covariates for score prediction in two steps: identify the top 200 covariates with highest prevalence and create indicator variables for each based on intensity (frequency: at least once, sporadic, frequent) prior to exposure relative to others in analytic sample. The recommendation is that these covariates are then prioritized (ranked) based on potential impact of confounding using quantities such as the Bross formula (Haris 2021 *arXiv*), and then selected for inclusion in the propensity model.

Identification of variable associated with the outcome and exposure, and not exposure alone is important to minimize instrumental variable bias (hdPS variables D in **Figure SM1**). Instrumental variable bias could increase bias and variance (decrease precision) of the estimated associations after propensity score adjustment. We aimed to minimize the inclusion of instrumental variables by performing variable selection prior to propensity score estimation.

**Figure SM1. Causal diagram for variable selection from empirically identified covariates**

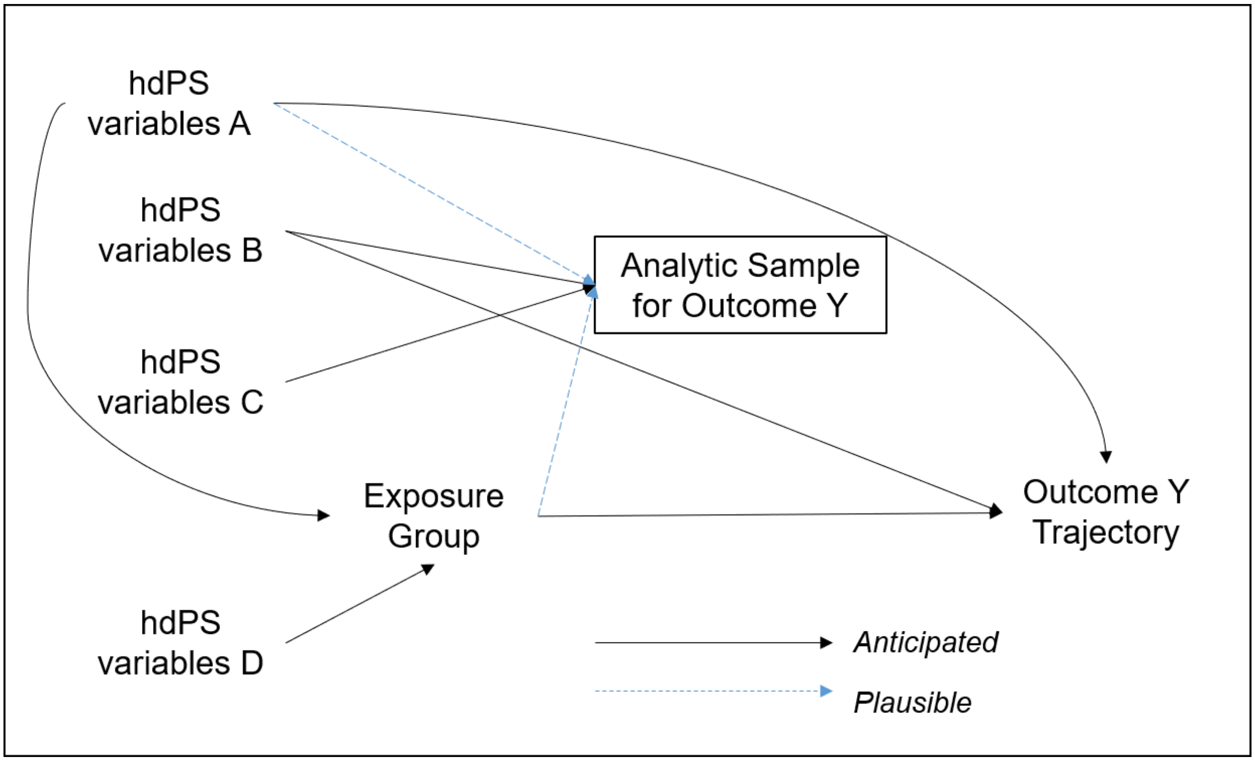

*Figure SM 1 Causal diagram for variable selection from empirically identified covariates*

Given the nature of outcomes in this study (longitudinal, irregular time intervals between measurements), we used linear mixed models fit using maximum likelihood for variable selection. We assessed the statistical significance of the model fit of each covariate indicator variable using likelihood ratio tests (df = 1), in addition to a model that includes only the time from origin date (index date + 30 days) as a continuous variable. We applied a false discovery rate adjusted p-value cutoff of 0.05 (Benjamini 1995 *J Royal Stat. Soc.*) to identify variables associated with the longitudinal outcome (**Table SM2**).

**Table SM2. Empirically identified covariates for the main analysis**

| Data Dimension | Variables Identified | Variables selected for categorization | HD covariates associated with longitudinal outcome # | HD covariates with at least 100 observations per exposure group | Additional variables associated with outcome availability during follow-up^ |
|---|---|---|---|---|---|
| Inpatient diagnoses | 537 | 200 | 387 | 6 | 0 out of 66 |
| Outpatient diagnoses | 537 | 200 | 512 | 438 | 64 out of 71 |
| Inpatient procedures | 1951 | 200 | 451 | 302 | 38 out of 142 |
| Outpatient procedures | 1951 | 200 | 499 | 409 | 64 out of 90 |
| Prescribing | 89 | 89 | 223 | 199 | 35 out of 36 |
| Labs | 877 | 200 | 312 | 231 | 54 out of 107 |
| **Total** | **5942** | **1089** | **2341** | **1585** | **253 out of 512** |

### hdPS variables A, B. ^ hdPS variables C

***Step 3. Propensity score models for exposure membership***

We restricted further analysis to high dimensional covariates with at least 100 observations within each exposure group. We followed a three-step process to estimate propensity scores (PS) for the study sample (n = 249,743). The objective is to develop a generalizable multiclass prediction model for PS that balances all pre-treatment covariates across the exposure groups (Exposed, Unexposed, Historical Control).

3.1 Model selection: We split the dataset (70/30) into training and test sets, stratified by exposure group. We used five-fold cross-validation for model selection with the highest area under the receiver operating characteristic curve (the macro-weighted AUC).

We used random forests algorithm (Brieman 2001 *Machine Learning*) fit using the ranger package (v 0.14.1) under four combinations of number of trees (number: 1000, 2000), minimum node size (size: 10, 25). The default number of variables sampled per split was floor(sqrt(number of columns)). Although overfitting PS models was identified as "not necessarily harmful" (Rosenbaum 1987 *JASA*), some authors suggest that this may lead to inflation of variance of estimates of association when applying them for confounding adjustment (Schuster 2016 *J Clin Epidemiol*). We included all investigator specified covariates and only those empirically identified covariates associated with any of the longitudinal outcomes. We used random forests with penalization given the algorithm inherently performs variable selection and incorporates statistical interactions between variables. Since socio-demographic characteristics were included in the prediction model, we did not fit separate prediction models when assessing subgroup differences.

The similar performance in the training and test sets for the propensity score models suggest that the model did not overfit. We assume that any positivity violation that the model may have encountered during model fitting were random, and not structural (Hernan and Robins 2023 *What If* Chapter 12).

3.2 Model fit: We fit the complete dataset using the most parsimonious set of parameters to predict the PS of membership in the exposure group which each person belongs to. We trimmed extreme propensity score weights to 2.5$^{th}$ percentile (low) and 97.5$^{th}$ percentile (high). We used the inverse of the propensity score as the inverse probability of treatment weight (IPTW).

3.3 Stabilized inverse probability of treatment weights (SIPTW): We multiplied the inverse probability of treatment weights with the prevalence of each outcome to obtain stabilized IPTW (Austin 2015 *Stat in Med*), which is beneficial for those with extreme IPTW (very low or very high probability of membership in an exposure group).

3.4 Covariate balance: We assessed covariate balance by plotting the distribution of the probability of membership in different cohorts.

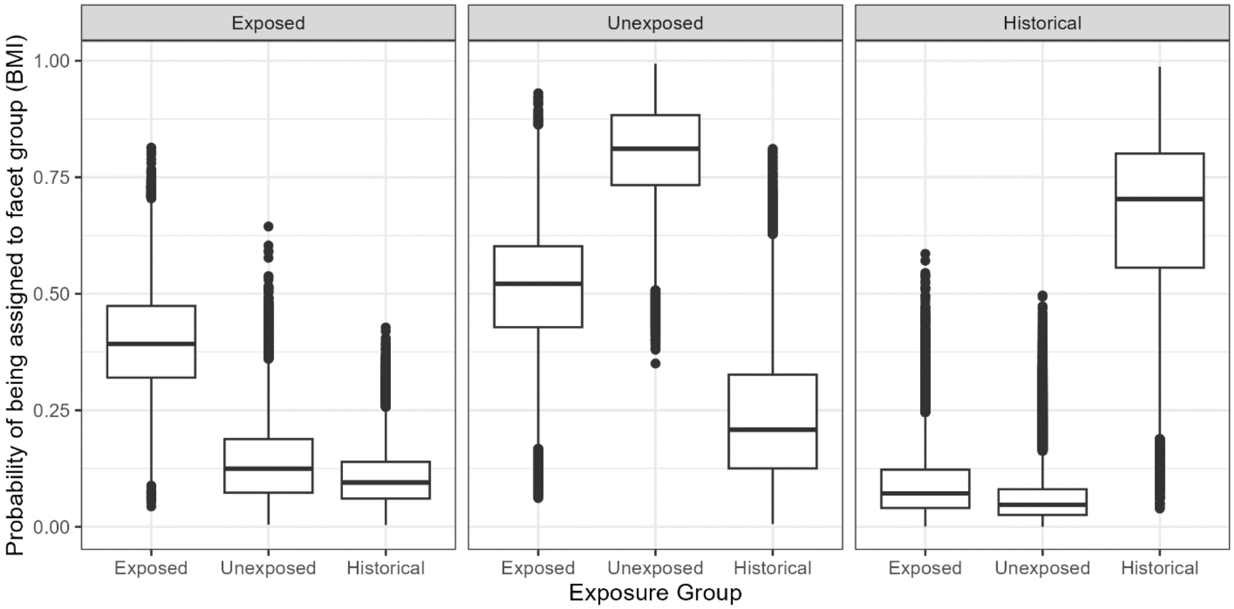

*Figure SM 2 Distribution of probability of membership in different cohorts*

We further examined population standardized bias for investigator specified covariates after weighting by SIPTW (McCaffrey 2013 *Stat in Med*). Population standardized bias assesses balance if the weighted mean/proportion of each exposure group differs relative to the pooled, unweighted mean/proportion of the study sample.

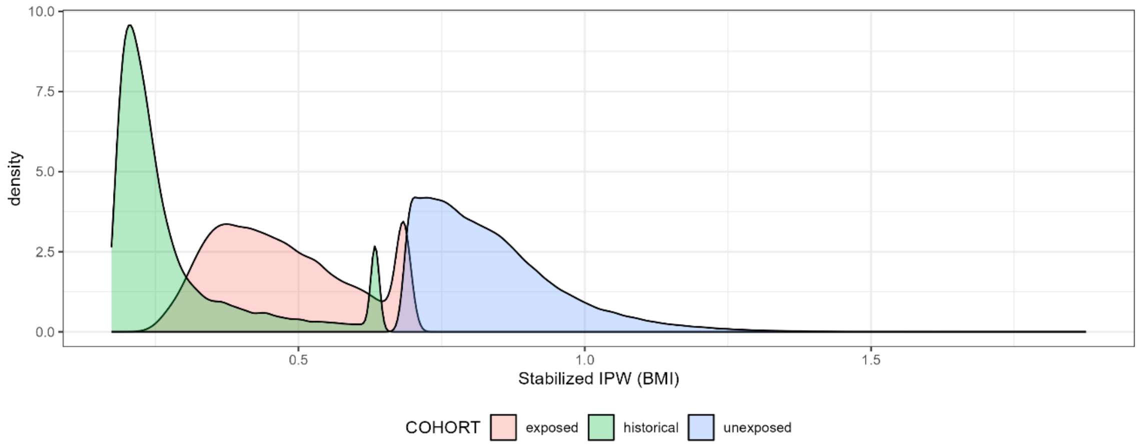

*Figure SM 3 Distribution of stabilized inverse probability weights after trimming*

##### *Step 4. Propensity score models for follow-up*

We split the dataset (70/30) into training and test sets, stratified by exposure group among patients with (n = 194,792 out of 249,743) and without BMI in the follow-up period. In the set of predictors, we additionally included the exposure group and variables associated with the follow-up status for that outcome, but not with the outcome trajectory for any outcome. We followed similar parameter tuning and performance metrics as Step 3.

Given the non-overlap in stabilized inverse probability of treatment weights, we additionally adjusted for all imbalanced investigator-specified covariates in the final outcome regression model.
