## Supplementary material for "Body mass index changes and their association with SARS-CoV-2 infection: a real-world analysis": RECORD-PE Checklist

The RECORD statement for pharmacoepidemiology (RECORD-PE) checklist of items, extended from the STROBE and RECORD statements, which should be reported in non-interventional pharmacoepidemiological studies using routinely collected health data

| Item No | STROBE items | RECORD items | RECORD-PE items | Page No |
| --- | --- | --- | --- | --- |
| **Title and abstract** | | | | |
| 1 | (a) Indicate the study’s design with a commonly used term in the title or the abstract.  (b) Provide in the abstract an informative and balanced summary of what was done and what was found. | 1.1: The type of data used should be specified in the title or abstract. When possible, the name of the databases used should be included.  1.2: If applicable, the geographical region and timeframe within which the study took place should be reported in the title or abstract.  1.3: If linkage between databases was conducted for the study, this should be clearly stated in the title or abstract. | — | 1, 4 |
| **Introduction** | | | | |
| Background rationale | | | | |
| 2 | Explain the scientific background and rationale for the investigation being reported. | — | — | 6 |
| Objectives | | | | |
| 3 | State specific objectives, including any prespecified hypotheses. | — | — | 6 |
| **Methods** | | | | |
| Study design | | | | |
| 4 | Present key elements of study design early in the paper. | — | 4.a: Include details of the specific study design (and its features) and report the use of multiple designs if used.  4.b: The use of a diagram(s) is recommended to illustrate key aspects of the study design(s), including exposure, washout, lag and observation periods, and covariate definitions as relevant. | 7 |
| Setting | | | | |
| 5 | Describe the setting, locations, and relevant dates, including periods of recruitment, exposure, follow-up, and data collection. | — | — | 7-8 |
| Participants | | | | |
| 6 | (a) Cohort study—give the eligibility criteria, and the sources and methods of selection of participants. Describe methods of follow-up. Case-control study—give the eligibility criteria, and the sources and methods of case ascertainment and control selection. Give the rationale for the choice of cases and controls. Cross sectional study—give the eligibility criteria, and the sources and methods of selection of participants.  (b) Cohort study—for matched studies, give matching criteria and number of exposed and unexposed. Case-control study—for matched studies, give matching criteria and the number of controls per case. | 6.1: The methods of study population selection (such as codes or algorithms used to identify participants) should be listed in detail. If this is not possible, an explanation should be provided.  6.2: Any validation studies of the codes or algorithms used to select the population should be referenced. If validation was conducted for this study and not published elsewhere, detailed methods and results should be provided.  6.3: If the study involved linkage of databases, consider use of a flow diagram or other graphical display to demonstrate the data linkage process, including the number of individuals with linked data at each stage. | 6.1.a: Describe the study entry criteria and the order in which these criteria were applied to identify the study population. Specify whether only users with a specific indication were included and whether patients were allowed to enter the study population once or if multiple entries were permitted. See explanatory document for guidance related to matched designs. | 7-8 |
| Variables | | | | |
| 7 | Clearly define all outcomes, exposures, predictors, potential confounders, and effect modifiers. Give diagnostic criteria, if applicable. | 7.1: A complete list of codes and algorithms used to classify exposures, outcomes, confounders, and effect modifiers should be provided. If these cannot be reported, an explanation should be provided. | 7.1.a: Describe how the drug exposure definition was developed.  7.1.b: Specify the data sources from which drug exposure information for individuals was obtained.  7.1.c: Describe the time window(s) during which an individual is considered exposed to the drug(s). The rationale for selecting a particular time window should be provided. The extent of potential left truncation or left censoring should be specified.  7.1.d: Justify how events are attributed to current, prior, ever, or cumulative drug exposure.  7.1.e: When examining drug dose and risk attribution, describe how current, historical or time on therapy are considered.  7.1.f: Use of any comparator groups should be outlined and justified.  7.1.g: Outline the approach used to handle individuals with more than one relevant drug exposure during the study period. | STable 1  8-10 |
| Data sources/measurement | | | | |
| 8 | For each variable of interest, give sources of data and details of methods of assessment (measurement). Describe comparability of assessment methods if there is more than one group. | — | 8.a: Describe the healthcare system and mechanisms for generating the drug exposure records. Specify the care setting in which the drug(s) of interest was prescribed. | 8-10 |
| Bias | | | | |
| 9 | Describe any efforts to address potential sources of bias. | — | — | 11-12 |
| Study size | | | | |
| 10 | Explain how the study size was arrived at. | — | — | SFig 2 |
| Quantitative variables | | | | |
| 11 | Explain how quantitative variables were handled in the analyses. If applicable, describe which groupings were chosen, and why. | — | — | 8-11 |
| Statistical methods | | | | |
| 12 | (a) Describe all statistical methods, including those used to control for confounding.  (b) Describe any methods used to examine subgroups and interactions.  (c) Explain how missing data were addressed.  (d) Cohort study—if applicable, explain how loss to follow-up was addressed. Case-control study—if applicable, explain how matching of cases and controls was addressed. Cross sectional study—if applicable, describe analytical methods taking account of sampling strategy.  (e) Describe any sensitivity analyses. | — | 12.1.a: Describe the methods used to evaluate whether the assumptions have been met.  12.1.b: Describe and justify the use of multiple designs, design features, or analytical approaches. | 11-13 |
| Data access and cleaning methods | | | | |
| 12 | — | 12.1: Authors should describe the extent to which the investigators had access to the database population used to create the study population.  12.2: Authors should provide information on the data cleaning methods used in the study. | — | 7 |
| Linkage | | | | |
| 12 | — | 12.3: State whether the study included person level, institutional level, or other data linkage across two or more databases. The methods of linkage and methods of linkage quality evaluation should be provided. | — | 7 |
| **Results** | | | | |
| Participants | | | | |
| 13 | (a) Report the numbers of individuals at each stage of the study (eg, numbers potentially eligible, examined for eligibility, confirmed eligible, included in the study, completing follow-up, and analysed).  (b) Give reasons for non-participation at each stage.  (c) Consider use of a flow diagram. | 13.1: Describe in detail the selection of the individuals included in the study (that is, study population selection) including filtering based on data quality, data availability, and linkage. The selection of included individuals can be described in the text or by means of the study flow diagram. | — | SFig 2 |
| Descriptive data | | | | |
| 14 | (a) Give characteristics of study participants (eg, demographic, clinical, social) and information on exposures and potential confounders.  (b) Indicate the number of participants with missing data for each variable of interest.  (c) Cohort study—summarise follow-up time (eg, average and total amount). | — | — | Table 1  13-14 |
| Outcome data | | | | |
| 15 | Cohort study—report numbers of outcome events or summary measures over time. Case-control study—report numbers in each exposure category, or summary measures of exposure. Cross sectional study—report numbers of outcome events or summary measures. | — | — | STable 4  Table 1  14 |
| Main results | | | | |
| 16 | (a) Give unadjusted estimates and, if applicable, confounder adjusted estimates and their precision (eg, 95% confidence intervals). Make clear which confounders were adjusted for and why they were included.  (b) Report category boundaries when continuous variables are categorised.  (c) If relevant, consider translating estimates of relative risk into absolute risk for a meaningful time period. | — | — | 14-15 |
| Other analyses | | | | |
| 17 | Report other analyses done—eg, analyses of subgroups and interactions, and sensitivity analyses. | — | — | 15 |
| **Discussion** | | | | |
| Key results | | | | |
| 18 | Summarise key results with reference to study objectives. | — | — | 16 |
| Limitations | | | | |
| 19 | Discuss limitations of the study, taking into account sources of potential bias or imprecision. Discuss both direction and magnitude of any potential bias. | 19.1: Discuss the implications of using data that were not created or collected to answer the specific research question(s). Include discussion of misclassification bias, unmeasured confounding, missing data, and changing eligibility over time, as they pertain to the study being reported. | 19.1.a: Describe the degree to which the chosen database(s) adequately captures the drug exposure(s) of interest. | 17 |
| Interpretation | | | | |
| 20 | Give a cautious overall interpretation of results considering objectives, limitations, multiplicity of analyses, results from similar studies, and other relevant evidence. | — | 20.a: Discuss the potential for confounding by indication, contraindication or disease severity or selection bias (healthy adherer/sick stopper) as alternative explanations for the study findings when relevant. **[A: Original text indicated this item was RECORD (ie, not RECORD-PE)?]** | 18 |
| Generalisability | | | | |
| 21 | Discuss the generalisability (external validity) of the study results. | — | — | 16-17 |
| **Other information** | | | | |
| Funding | | | | |
| 22 | Give the source of funding and the role of the funders for the present study and, if applicable, for the original study on which the present article is based. | — | — | 1 |
| Accessibility of protocol, raw data, and programming code | | | | |
| 22 | — | 22.1: Authors should provide information on how to access any supplemental information such as the study protocol, raw data, or programming code. | — | 19 |

RECORD=reporting of studies conducted using observational routinely collected data; RECORD-PE=RECORD for pharmacoepidemiological research; STROBE=strengthening the reporting of observational studies in epidemiology.

*REFERENCE: Langan SM, Schmidt S, Wing K, Ehrenstein V, Nicholls S, Filion K, Klungel O, Petersen I, Sorensen H, Guttmann A, Harron K, Hemkens L, Moher D, Schneeweiss S, Smeeth L, Sturkenboom M, von Elm E, Wang S, Benchimol EI. The REporting of studies Conducted using Observational Routinely-collected health Data (RECORD) Statement for Pharmacoepidemiology (RECORD-PE). *BMJ* 2018; 363: k3532.
